# TRansit ACessibility Tool (TRACT): Developing a novel scoring system for public transportation system accessibility

**DOI:** 10.1101/2023.03.07.23286932

**Authors:** Erica Twardzik, Jennifer Schrack, Keshia M. Pollack Porter, Taylor Coleman, Kathryn Washington, Bonnielin K. Swenor

## Abstract

**Introduction:** Although federal laws require equal access to public transportation for people with disabilities, access barriers persist. Lack of sharing accessibility information on public transportation websites restricts people with disabilities from making transportation plans and effectively using public transportation systems. This project aims to document information provided about public transportation systems accessibility and share this information using an open data platform.

**Methods:** We reviewed the top twenty-six public transportation systems in the United States based on federal funding in fiscal year 2020. Information about accessibility was abstracted from the webpages of each public transportation system by two independent reviewers from February-March 2022. Informed by universal design principles, public transportation systems were scored across six dimensions: facility accessibility (0-22 points), vehicle accessibility (0-11 points), inclusive policies (0-12 points), rider accommodations (0-9 points), paratransit services (0-6 points), and website accessibility (0-2 points). Total scores were calculated as the sum of each dimension and ranged from 0-62 points. Data and findings were publicly disseminated (https://disabilityhealth.jhu.edu/transitdashboard/).

**Results:** The average overall accessibility information score was 31.9 (SD=6.2) out of 62 possible points. Mean scores were 8.4 (SD=2.9) for facility accessibility, 4.5 (SD=2.1) for vehicle accessibility, 7.8 (SD=1.6) for inclusive policies, 4.9 (SD=1.6) for rider accommodations, 4.5 (SD=2.0) for paratransit services, and 1.8 (SD=0.4) for website accessibility. Eleven public transportation systems (42%) received the maximum score for paratransit services and 20 (77%) received the maximum score for website accessibility. No public transportation system received the maximum score for any of the other dimensions.

**Conclusions:** Using a novel scoring system, we found significant variation in the accessibility information presented on public transportation system websites. Websites are a primary mode where users obtain objective information about public transportation systems and are therefore important platforms for communication. Absence of accessibility information creates barriers for the disability community and restricts equal access to public transportation.

## 1. Introduction

Although laws have been adopted to protect the rights of people with disabilities onboard public transportation systems, the disability community continues to face barriers accessing public transportation.^5^ For example, people with disabilities encounter uneven boarding surfaces, gaps between vehicles and platforms, inoperable elevators, and inaccessible curbs when trying to access public transportation.^5^ Insurmountable barriers within the public transportation environment often force people with disabilities to go out of their way to find alternative routes, and make travel prohibitive. A greater proportion of people with disabilities live in zero-vehicle households compared to people without disabilities, leading to increased need for public transportation access among people with disabilities.^6^ Subsequently, inaccessible public transportation disproportionately affects people with disabilities. These characteristics make accessible transportation a key social determinant of health and a driver of health inequities among people with disabilites.^7, 8^

Public transportation is not only a critical social determinant of health, but also indirectly impacts almost every other aspect of health by either providing or hindering access to services and destinations. Barriers to accessible transportation for people with disabilities create addressable, inequitable access to food,^9^ education,^10^ health care,^11, 12^ social participation,^10^ and employment.^13^ Although the disability community has repeatedly identified barriers to accessing public transportation across the U.S.^14, 15^, there is a lack of national data identifying and tracking public transportation accessibility. Information about how public transportation accessibility is communicated to the public is needed to document systemic barriers people with disabilities face to make plans and use public transportation.

The Federal Transit Administration (FTA), responsible for ensuring compliance within the Americans with Disabilities Act (ADA), collects comprehensive assessments of public transportation system. An FTA assessment includes stakeholder interviews, meetings with public transportation personnel, and an on-site evaluation. However, there are limitations to this model. Specifically, the FTA is responsible for overseeing over 2,000 transportation systems and assessments of ADA compliance only occur in response to complaints from users. Between 2000 and 2017 the FTA evaluated 31 fixed-route operations (<2% of transportation systems) and has summarized the results within 33 publicly available reports.^16^ The FTA evaluations are resource intensive (e.g., time, cost) which limits the feasibility of conducting annual evaluations across all public transit systems.

Public transportation websites are a primary mode through which users obtain information about public transportation systems^23^ and information gathering is the first step in a travel journey chain.^24^ Documenting and comparing the accessibility information on public transportation system websites presents an opportunity to begin to develop a surveillance system that identifies targets for future policy and practice interventions promoting transportation equity among people with disabilities. This project aims to document and examine publicly available information about public transportation accessibility available on system websites.

## 2. Material and methods

### 2.1 TRansit ACessibility Tool (TRACT) Development

The TRACT was developed by reviewing current literature, integrating universal design principles, discussing constructs with people with disabilities, and consulting subject matter experts. This work was guided by universal design principles, which are a useful guide to understand the usability of public transportation systems.^17, 18^ Rather than modifying a system to accommodate a specific individual,^19^ universal design aims to design environments to be accessible for all people, including people with disabilities. Guided by seven core principles, universal design aims to design systems and spaces that are accessible, usable, and inclusive.^20^ The TRACT aims to identify accessibility information on public transportation system websites that promotes usability, accessibility, and inclusion. Numerous items included within TRACT were informed by resources from the Victoria Transport Policy Institute, the American Public Transportation Association, and the National Aging and Disability Transportation Center.^21, 25, 26^ Using an iterative process, our research team initially refined and pilot tested the TRACT on ten Canadian public transportation systems. Additional items were added to the TRACT during the testing phase. Coding of the final four public transportation systems of the testing phase revealed no new features suggesting that saturation of items within TRACT was satisfactory. All items collected from public transportation systems are listed within **Appendix B**.

### 2.2 Data Collection Procedures

Using a novel, universal design-informed scoring system, we abstracted and compared data across key accessibility features of public transportation. Two researchers were trained to code the TRACT between January and February of 2022. Training consisted of instructional videos, close reading of the manual, and virtual meetings to review individual TRACT items. During virtual meetings each item was discussed in a group setting using written, visual, and verbal instructions. Both coders were oriented to the digital TRACT form hosted by Qualtrics (Qualtrics, Provo, UT), an online web application for building and managing online surveys and databases. Practice public transportation systems were independently coded and then discussed within a group setting until there was consensus among all coders. To be certified to code independently, five public transportation system websites needed to be coded with inter-rater reliability ≥ 85%.

From February through March of 2022, three trained researchers (ET, TC, and KW) qualitatively coded universal design features advertised on public transportation websites. Within this pilot project, the top twenty-six public transportation systems in the US receiving federal funding in 2020 were examined (**Table 1**).^27^ Public transportation systems receiving the highest dollar amount of total federal funding in fiscal year 2020 were identified using the National Transit Database rankings.^27^ The total federal funds allocated to a public transportation system is the sum of all Federal Transit Administration funds, U.S. Department of Transportation funds, and funding from departments of the federal government other than transportation during fiscal year 2020. The publicly available website landing page for each public transportation system was identified through a web search of the public transportation system name. In the case where multiple public transportation systems shared one landing page (e.g., MTA New York City Transit and MTA Long Island Railroad), the public transportation stystems were grouped together as one public transportation system.

**Table 1.** Accessible public transportation system dimensions, number of items, example items, and relevance.

| Dimensions of Transportation Accessibility | # of Items | Example items | Relevance |
| --- | --- | --- | --- |
| Facility accessibility (range 0-22) | 132 | Bus stop wayfinding signs, train station audio and visual announcements | Accessible physical environments of bus stops or train stations are required for inclusion. |
| Vehicle accessibility (range 0-11) | 66 | Ramps, lifts, securement devices | Accessible physical environments of buses or trains are necessary to board and use the service. |
| Inclusive policies (range 0-12) | 12 | Public involvement | Users, including people with disabilities, need to be involved during the public transportation planning process. |
| Rider accommodations (range 0-9) | 14 | Text telephone available to submit accommodation request | Modes through which accommodation requests are submitted need to be accessible to all users. |
| Paratransit services (range 0-6) | 33 | Eligibility determination, hours of operation | Limitations in eligibility or operation of paratransit service restricts access for people with disabilities. |
| Website accessibility (range 0-2) | 6 | Alternative text, webpage contrast | Website environments are a primary platform for users to obtain information about transportation systems. |

Researchers individually recorded information using a Qualtrics survey prior to weekly consensus meetings. During weekly meetings, researchers discussed any coding disagreement to reach a consensus for each public transportation system. Consensus codes represented the final decision for each item within the public transportation system and were recorded within a new Qualtrics form. Within each public transportation system website landing page, coders documented category or subcategory websites with information about the public transportation system accessibility, transit maps, real-time information, web-based form to submit an accommodation request, and web-based form to submit a complaint. Therefore, a maximum of six unique web addresses were documented for each public transportation system.

Website accessibility data collection among the public transportation system websites was carried out using previously described methods.^28^ Briefly, website accessibility was assessed using a combination of automatic and high-level manual accessibility testing. Automatic accessibility data was collected using the WAVE^®^ accessibility tool.^29^ The WAVE^®^ accessibility tool generates accessibility information scores based on the number of detected errors, density of errors, and number of likely or potential accessibility errors. These scores are then normalized to the WebAIM Million sample of one million homepages in March 2022 to generate an automatic accessibility information score.^30^ Additionally, high-level manual accessibility testing was carried out by expert testers who evaluated multiple aspects of web accessibility (e.g., accuracy of defined language, appropriate of image alternative text, low contrast content) on the public transportation system landing webpage, accessibility webpage, transit maps webpage, and one random selected webpage from the remaining three webpages (i.e., real-time information, accommodation request, complaint). The manual test results generated an accessibility information score which was averaged with the automatic accessibility information score to produce an overall site accessibility information score.

### 2.3 Scoring TRACT Data

Using inductive reasoning, dimensions of accessibility were created by clustering together individual items. The six dimensions of public transportation system accessibility included: facility accessibility, vehicle accessibility, inclusive policies, rider accommodations, paratransit services, and website accessibility.^17, 18^ Facility accessibility included information about the accessibility of the stops or stations. Items within the facility accessibility dimension included wayfinding signs and audio/visual announcements present at pick-up and drop-off locations (e.g., bus stop, train station). Vehicle accessibility information is critical for users to know if they can board and use the public transportation service. Items within the vehicle accessibility dimension included items on the transportation vehicle (e.g., buses, trains) such as ramps, lifts, and securement devices. Inclusive policies represent accessibility policies and ongoing efforts to consult public transportation users during the planning and prioritizing process the public transportation system improvements. Rider accommodations include modes through which users can request accommodations, and if the modes are accessible to people with disabilities. Paratransit services includes information about the hours of operation of paratransit and if the paratransit system is transparent about eligibility determination for using the service. Website accessibility is critical for public transportation accessibility because it is the primary platform through which users obtain information about the public transportation system. Website accessibility included features such as alternative text and webpage contrast, features that are necessary for equal access to information about the public transportation system. **Table 1** summarizes the score range, number of items, example items, and relevance for accessible public transportation for each dimension.

Item level recoding, dimension creation, and overall accessibility information score generation is detailed in **Appendix C**. The facility accessibility and vehicle accessibility dimensions include information across multiple different modes of public transportation (i.e., bus, light rail, heavy rail, paratransit). Therefore, each accessibility feature was averaged across available modes for each transportation system and then summed to create a total score. Facility accessibility was comprised of 132 items that were summarized into a score ranging from 0 to 22. Vehicle accessibility was comprised of 66 items, which were summarized into a score ranging from 0 to 11. The inclusive policies dimension was calculated by summing twelve individual items. This resulted in an inclusive policies score ranging from 0 to 12. The rider accommodations dimension included fourteen items, with six items asking the same information across multiple different modes. Therefore, the six items were averaged across available modes for each transportation system. The remaining eight items plus the average of six items were summed to create a rider accommodations score ranging from 0 to 9. Information about paratransit services was captured with 33 items across the whole public transportation system. Fine grain detail on paratransit hours of operation were captured and summarized into tertiles of availability. A paratransit services score was calculated, ranging from 0 to 6. Lastly, the overall website accessibility values for a public transportation system were calculated as an average of the automated testing score and manual testing scores. The website accessibility information score was generated based on tertiles of the overall website accessibility values. The lowest tertile (overall accessibility values less than or equal to 3.3) received 0 points, the middle tertile (overall accessibility values greater than 3.3 and less than or equal to 6.7) received 1 point, and the highest tertile (overall accessibility values greater than 6.7) received 2 points.

The six dimensions described above summarize accessibility features communicated by public transportation systems on their publicly available websites. The overall accessibility information score is a summation of the dimension scores. Summary statistics were calcuated for the overall accessibility information score, as well as each dimension score (facility accessiblity, vehicle accessiblity, inclusive policies, rider accommodations, paratransit services, and website accessibility). All analyses were conducted using STATA 16.1. (College Station, TX) and syntax is provided in **Appendix D**.

Raw data, summary statistics, and figures were disseminated to the public on the Johns Hopkins Disability and Health Research Center Public Transit Disability Dasbhoard (https://disabilityhealth.jhu.edu/transitdashboard/).

### 2.4 Inter-Rater Reliability

For each individual TRACT item (e.g., travel training program), the number of public transit systems with the item present divided by the total number of public transit systems was calculated. This proportion represents the average prevalence of each TRACT item within the study sample. Percent agreement was calculated by summing instances when two coders selected the same response option (e.g., both coders agreed the item was present or both coders agreed the item was absent) divided by the total number of public transportation systems assessed. Inter-rater reliability was calculated using intra-class correlation and kappa statistics among continuous and categorical variables, respectively. Categorical items with a prevalence below 0.10 were excluded from the reliability analysis given the poor performance of Cohen’s kappa when prevalence of an item is extremely low.^31, 32^

## 3. Results

**Table 2** includes a complete list of public transit system characteristics (i.e., name, city, state), total federal funding received in fiscal year 2020, and the website landing page among the top twenty-six public transportation systems evaluated. Most public transportation systems had a unique website landing page to communicate information to users. However, the New York Metropolitan Transit Authority (NYMTA) hosted information for three public transportation systems (i.e., MTA New York City Transit, MTA Long Island Rail Road, MTA Metro-North Railroad) and was therefore categorized under one public transportation system. The amount of federal funds received in 2020 ranged from $3.8 billion to $154 million among the 26 public transportation systems evaluated as part of this project.

**Table 2.**
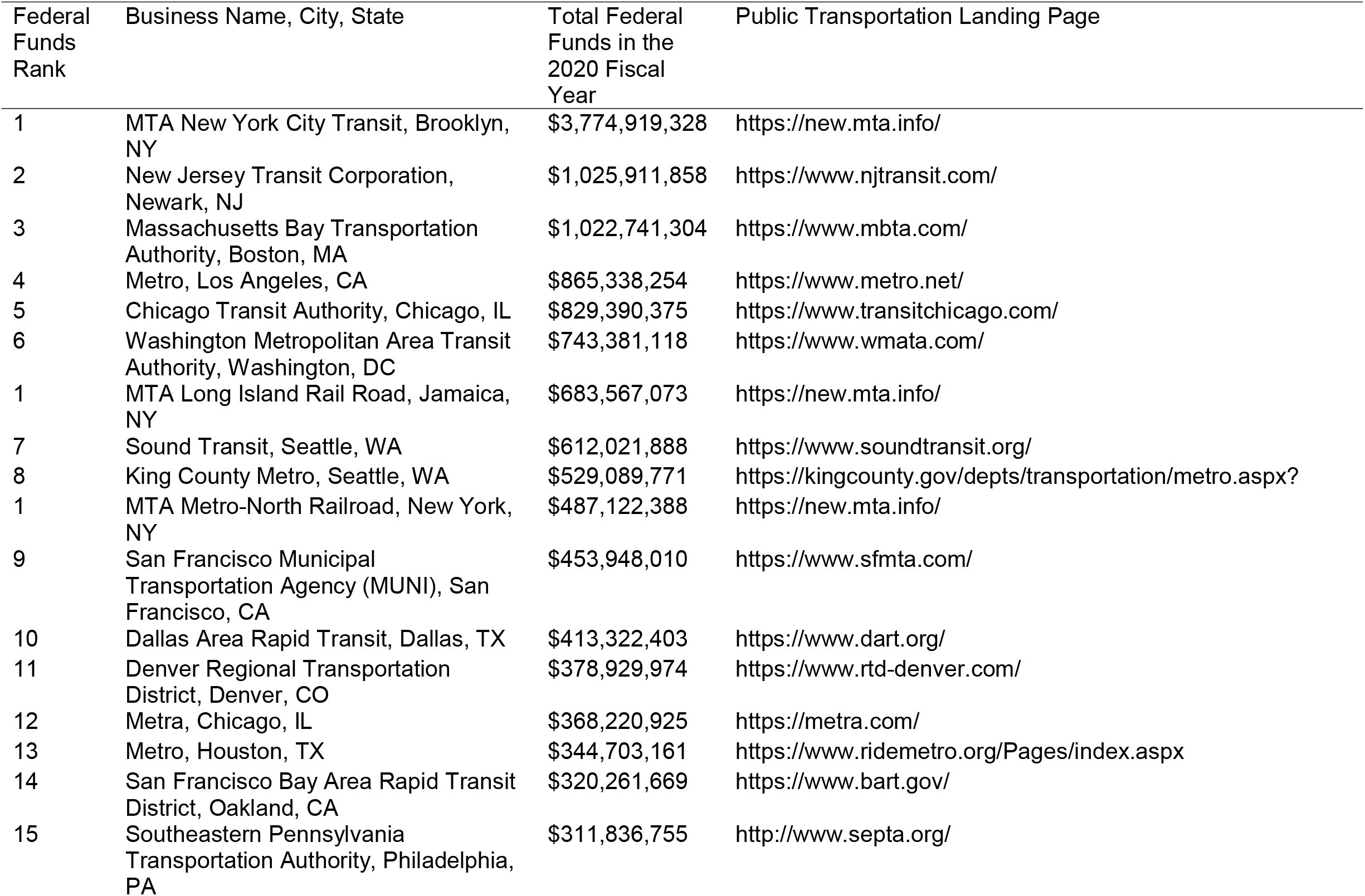
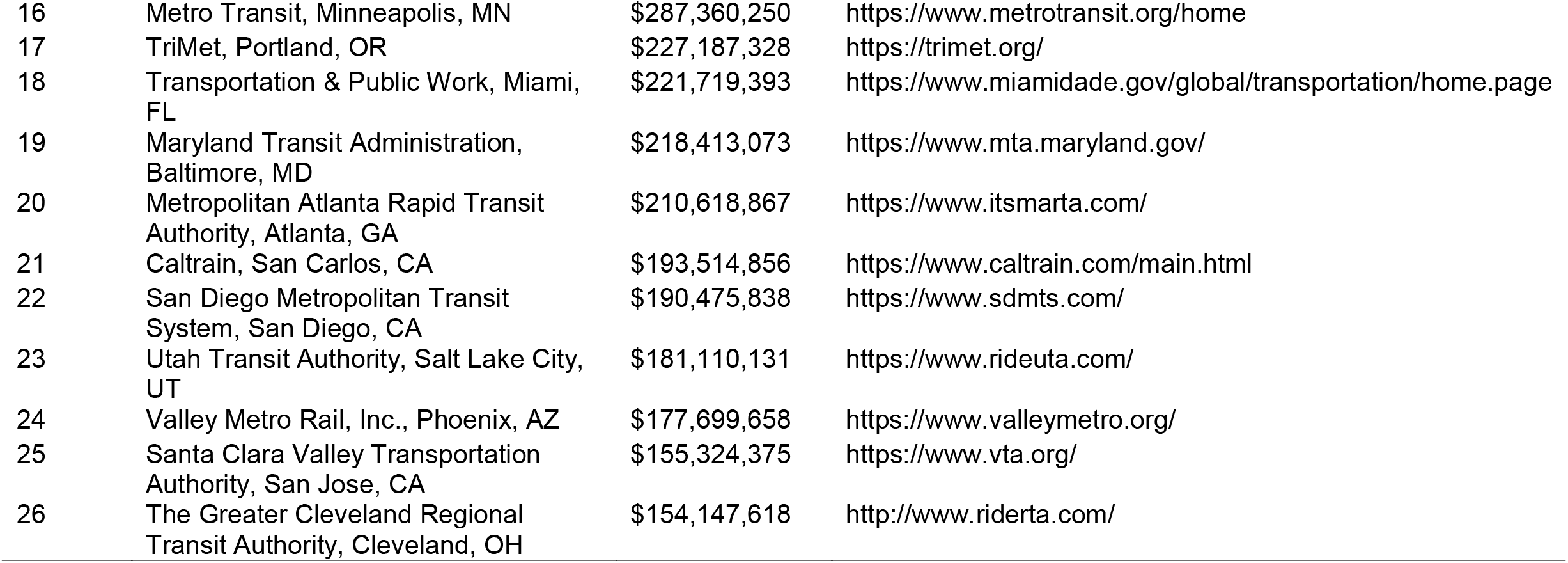
Public transportation agency characteristics (i.e., business name, city, state), total federal funding received, and website landing page among the top twenty-six public transportation systems evaluated in this study.

| Federal Funds Rank | Business Name, City, State | Total Federal Funds in the 2020 Fiscal Year | Public Transportation Landing Page |
| --- | --- | --- | --- |
| 1 | MTA New York City Transit, Brooklyn, NY | \$3,774,919,328 | https://new.mta.info/ |
| 2 | New Jersey Transit Corporation, Newark, NJ | \$1,025,911,858 | https://www.njtransit.com/ |
| 3 | Massachusetts Bay Transportation Authority, Boston, MA | \$1,022,741,304 | https://www.mbta.com/ |
| 4 | Metro, Los Angeles, CA | \$865,338,254 | https://www.metro.net/ |
| 5 | Chicago Transit Authority, Chicago, IL | \$829,390,375 | https://www.transitchicago.com/ |
| 6 | Washington Metropolitan Area Transit Authority, Washington, DC | \$743,381,118 | https://www.wmata.com/ |
| 1 | MTA Long Island Rail Road, Jamaica, NY | \$683,567,073 | https://new.mta.info/ |
| 7 | Sound Transit, Seattle, WA | \$612,021,888 | https://www.soundtransit.org/ |
| 8 | King County Metro, Seattle, WA | \$529,089,771 | https://kingcounty.gov/depts/transportation/metro.aspx? |
| 1 | MTA Metro-North Railroad, New York, NY | \$487,122,388 | https://new.mta.info/ |
| 9 | San Francisco Municipal Transportation Agency (MUNI), San Francisco, CA | \$453,948,010 | https://www.sfmta.com/ |
| 10 | Dallas Area Rapid Transit, Dallas, TX | \$413,322,403 | https://www.dart.org/ |
| 11 | Denver Regional Transportation District, Denver, CO | \$378,929,974 | https://www.rtd-denver.com/ |
| 12 | Metra, Chicago, IL | \$368,220,925 | https://metra.com/ |
| 13 | Metro, Houston, TX | \$344,703,161 | https://www.ridemetro.org/Pages/index.aspx |
| 14 | San Francisco Bay Area Rapid Transit District, Oakland, CA | \$320,261,669 | https://www.bart.gov/ |
| 15 | Southeastern Pennsylvania Transportation Authority, Philadelphia, PA | \$311,836,755 | http://www.septa.org/ |
| 16 | Metro Transit, Minneapolis, MN | \$287,360,250 | https://www.metrotransit.org/home |
| 17 | TriMet, Portland, OR | \$227,187,328 | https://trimet.org/ |
| 18 | Transportation & Public Work, Miami, FL | \$221,719,393 | https://www.miamidade.gov/global/transportation/home.page |
| 19 | Maryland Transit Administration, Baltimore, MD | \$218,413,073 | https://www.mta.maryland.gov/ |
| 20 | Metropolitan Atlanta Rapid Transit Authority, Atlanta, GA | \$210,618,867 | https://www.itSMARTA.com/ |
| 21 | Caltrain, San Carlos, CA | \$193,514,856 | https://www.caltrain.com/main.html |
| 22 | San Diego Metropolitan Transit System, San Diego, CA | \$190,475,838 | https://www.sdmts.com/ |
| 23 | Utah Transit Authority, Salt Lake City, UT | \$181,110,131 | https://www.rideuta.com/ |
| 24 | Valley Metro Rail, Inc., Phoenix, AZ | \$177,699,658 | https://www.valleymetro.org/ |
| 25 | Santa Clara Valley Transportation Authority, San Jose, CA | \$155,324,375 | https://www.vta.org/ |
| 26 | The Greater Cleveland Regional Transit Authority, Cleveland, OH | \$154,147,618 | http://www.riderta.com/ |

Among the 26 public transportation systems assessed, the mean total accessibility information score was 31.9 (SD=6.2) out of 62 possible total points and ranged from 16.3 to 40.7 (**Table 3**). The mean scores were 8.4 (SD=2.9) out of 22 points for facility accessibility, 4.5 (SD=2.1) out of 11 points for vehicle accessibility, 7.8 (SD=1.6) out of 12 points for inclusive policies, 4.9 (SD=1.6) out of 9 points for rider accommodations, 4.5 (SD=2.0) out of 6 points for paratransit services, and 1.8 (SD=0.4) out of 2 points for website accessibility. **Figure 1** displays a horizontal stacked bar chart displays the overall accessibility of the 26 public transportation systems assessed. The total bar length represents overall accessibility and the colors embedded within each horizontal bar represent the six dimensions of accessibility. Within our sample, Central Puget Sound Regional Transit Authority had the highest total score of 40.7 points and Transportation & Public Work of Miami-Dade had the lowest score with a total score of 16.3 points. Horizontal bar charts for each dimension are presented in **Figures A.1-A.6**, where the x-axis represents the theoretical range of each dimension, and a vertical line is drawn at half of the maximum points for each dimension. Eleven public transportation systems (42%) earned the maximum score for paratransit services and 20 (77%) earned the maximum score for website accessibility. No public transportation system earned the maximum score for facility accessibility, vehicle accessibility, inclusive policies, or rider accommodations. Dimensions where public transportation systems earned at least half of the maximum points included: 3 (12%) in facility accessibility, 11 (42%) in vehicle accessibility, 24 (92%) in inclusive policies, 16 (62%) in rider accommodations, 22 (85%) in paratransit services, and 26 (100%) in website accessibility.

**Table 3.**
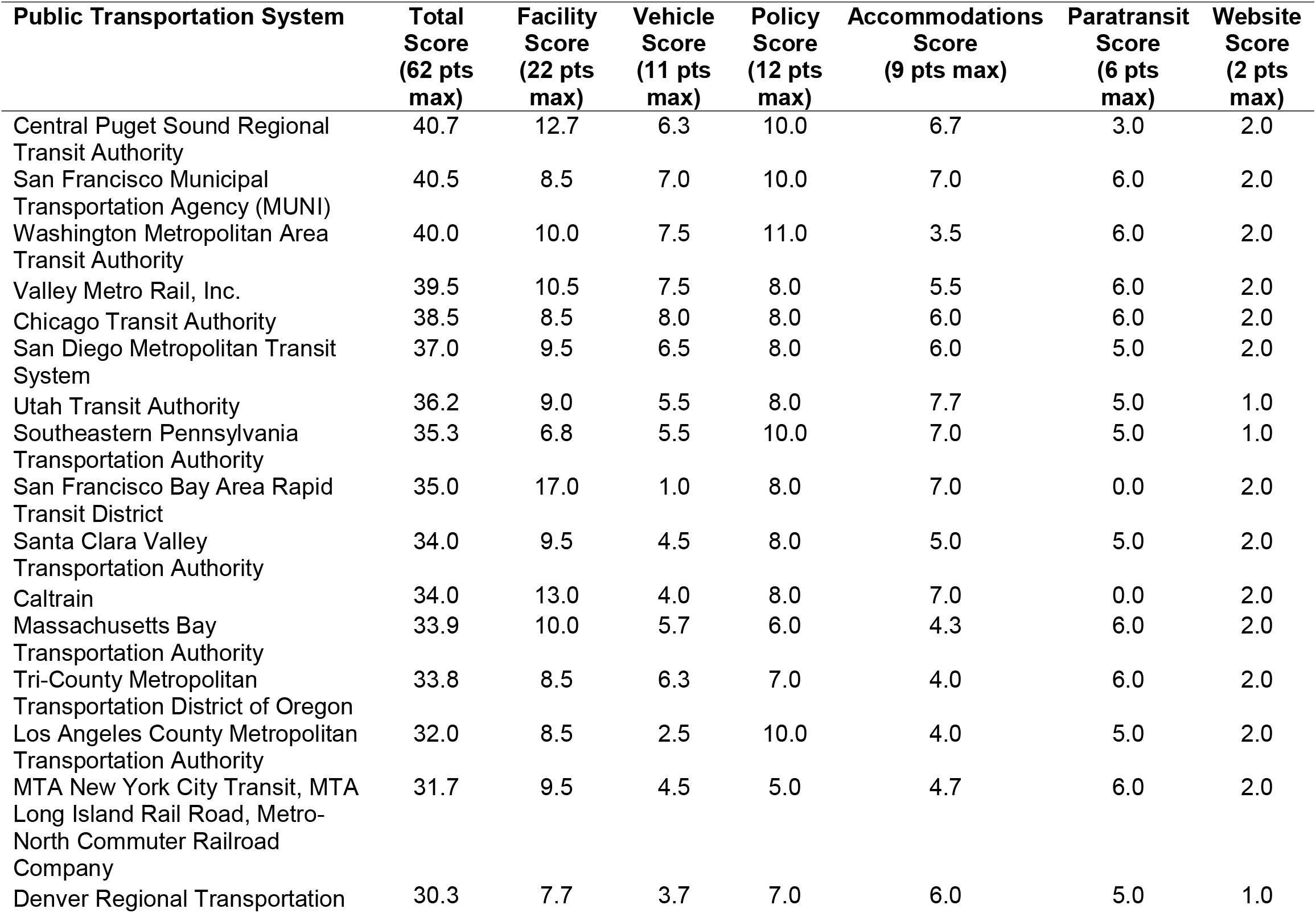
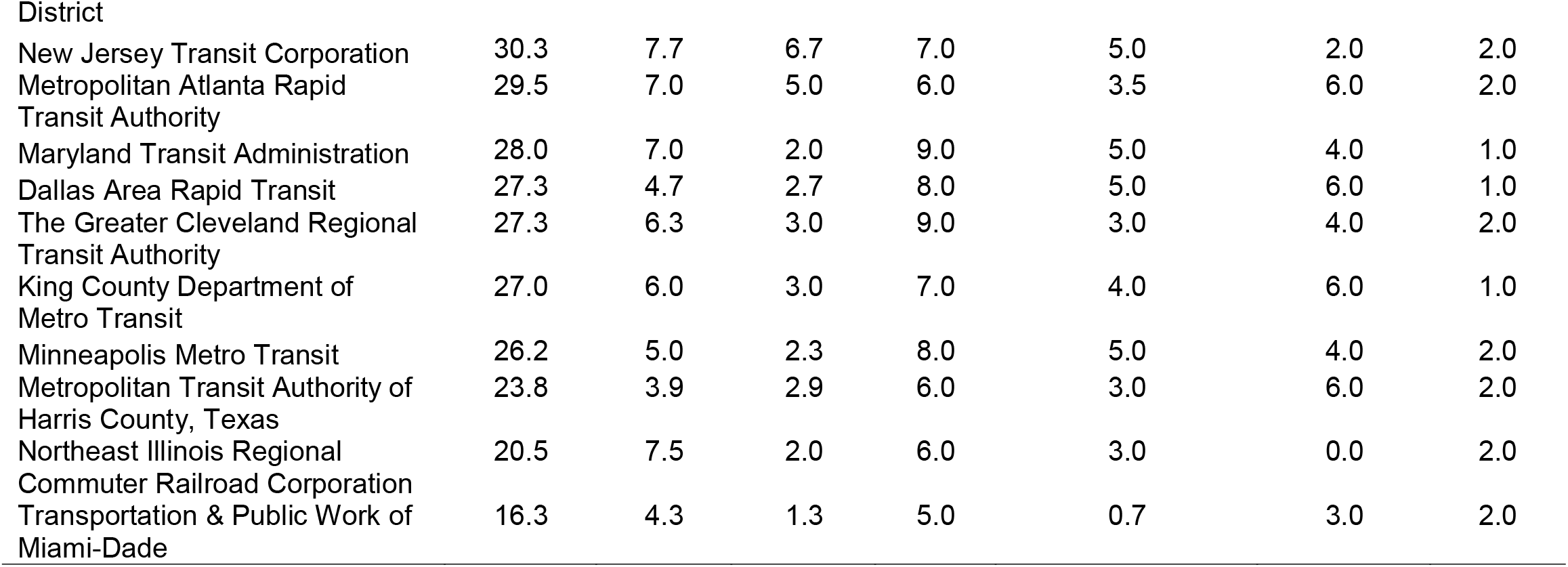
TRansit ACcessibility Tool (TRACT) scores for accessibility and disability inclusion among the top 26 public transportation systems receiving federal funding in the United States.

**Figure 1.**
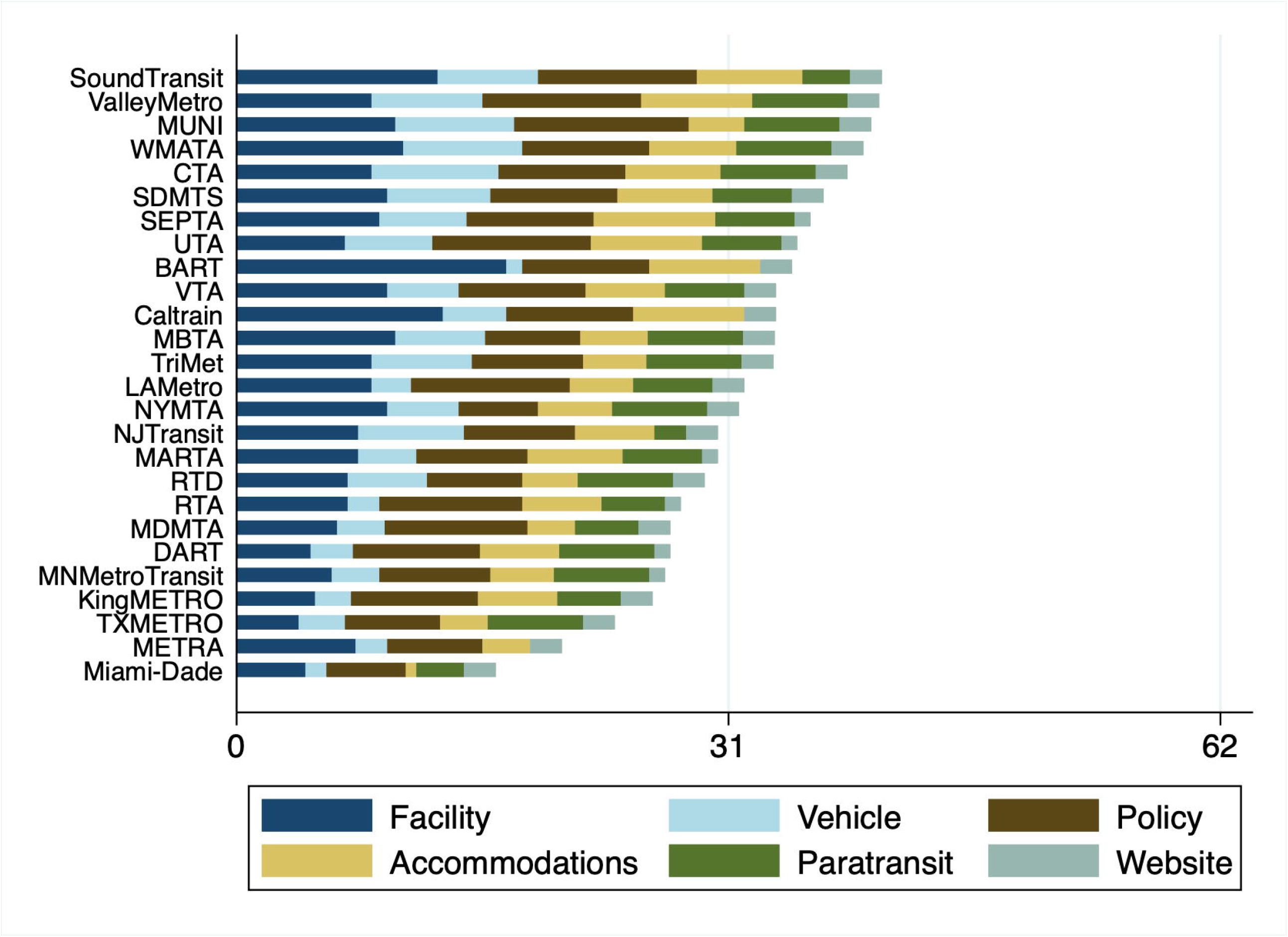
TRansit ACcessibility Tool (TRACT) scores for overall accessibility and disability inclusion among the top 26 public transportation systems receiving federal funding in the United States. *Note:* In order of appearance: SoundTransit = Central Puget Sound Regional Transit Authority; ValleyMetro = Valley Metro Rail, Inc.; MUNI = San Francisco Municipal Transportation Agency; WMATA = Washington Metropolitan Area Transit Authority; CTA = Chicago Transit Authority; SDMTS = San Diego Metropolitan Transit System; SEPTA = Southeastern Pennsylvania Transportation Authority; UTA = Utah Transit Authority; BART = San Francisco Bay Area Rapid Transit District; VTA = Santa Clara Valley Transportation Authority; MBTA = Massachusetts Bay Transportation Authority; TriMet = Tri-County Metropolitan Transportation District of Oregon; LAMetro = Los Angeles County Metropolitan Transportation Authority; NYMTA = Metropolitan Transit Authority New York City Transit, Metropolitan Transit Authority Long Island Rail Road, Metro-North Commuter Railroad Company; NJTransit = New Jersey Transit Corporation; MARTA = Metropolitan Atlanta Rapid Transit Authority; RTD = Denver Regional Transportation District; RTA = The Greater Cleveland Regional Transit Authority; MDMTA = Maryland Transit Administration; DART = Dallas Area Rapid Transit; MNMetroTransit = Minneapolis Minnesota Metro Transit; KingMETRO = King County Department of Metro Transit; TXMETRO = Metropolitan Transit Authority of Harris County, Texas; METRA = Northeast Illinois Regional Commuter Railroad Corporation; Miami-Dade = Transportation & Public Work of Miami-Dade

We observed moderate reliability (ICC = 0.47; 95% CI = -0.05, 0.76) in the overall score of transportation accessibility among the 26 public transportation systems assessed. Among the dimensions of public transportation accessibility, accommodations (ICC = 0.70; 95% CI = 0.43, 0.85) and paratransit services (ICC = 0.63; 95% CI = 0.33, 0.82) had substantial agreement between coders, vehicle accessibility (ICC = 0.51; 95% CI = -0.02, 0.78) had moderate agreement, inclusive policies (ICC = 0.30; 95% CI = -0.08, 0.61) had fair agreement, and facility accessibility (ICC = 0.17; 95% CI = -0.10, 0.47) had slight agreement (**Table E.1**). Reliability of website accessibility was not calculated due to the robust measurement combining both automatic and high-level manual accessibility testing. Some items used to calculate the six dimensions of public transportation accessibility were only relevant if another item was present. For example, level boarding of a bus system was only coded if a public transportation system included buses as a mode offered within their system. After considering skip logic, sample sizes ranged from 2 to 26. A total of 140 of 263 items had a prevalence greater than 0.1, and reliability statistics were calculated among the 140 items. Overall, 28 out of 140 items coded had Cohen’s kappa values in the substantial to almost perfect reliability range (0.61 to 1.0), 71 items had kappa values in the fair to moderate reliability range (0.21 to 0.60), and 41 items had kappa values within the poor to slight reliability range (less than 0.20). Detailed reliability statistics for each individual item are summarized within **Table E.2**.

## 4. Discussion

The TRACT novel scoring system identified correctable gaps in information about public transportation accessibility available to system websites. The results indicate that there is significant variation in information about public transportation accessibility across the twenty-six systems assessed, meaning the ability for people with disabilities to make plans and use public transportation depends on where these riders live. Furthermore, there was variation in information provided across six dimensions of accessibility, indicating where public transportation systems can make improvements in making accessibility information more available. These data suggest that every system evaluated has room for improvement, as no public transportation system received the maximum score for facility accessibility, vehicle accessibility, or rider accommodations.

Our findings add to the previous literature investigating barriers to public transportation for people with disabilities. Previous research has detailed experiences of people with disabilities using public transportation, and highlighted the numerous barriers encountered when attempting to use public transit.^33-35^ Barriers to public transportation use can be present at every stage of the travel chain including when clients are gathering information about the system, waiting at stops or stations, boarding and exiting the vehicle, and spending time inside the vehicle.^36^ Public transportation websites are a primary mode to obtain objective information about public transportation systems and are therefore important for information access. This project adds to current literature by highlighting a systemic issue with the communication of accessibility features on public transportation system websites in the United States and begins to establish a method to surveil these issues nationally. Previous research has been limited in its focus on a specific functional impairment, rather than integrating universal design features which would be relevant to all people with disabilities.^19^

### 4.1 Strengths and limitations

This is among the first studies to systematically evaluate the public transportation accessibility information. The TRACT scoring system is an efficient and low-cost method for evaluating the accessibility of public transportation systems within the United States. In contrast to FTA evaluations, the TRACT method provides a feasible approach for ongoing surveillance of public transportation accessibility throughout the United States. TRACT leverages information accessibility information communicated on public transportation websites to estimate accessibility of a public transportation system. This systematic method for capturing national accessibility information about public transportation systems can be used to evaluate current and future accessibility of public transportation systems. While FTA evaluations are critical to resolve complaints issued by an end user about public transportation system barriers, these evaluations are not systematically completed across public transportation systems at regular time intervals. While the American Communities Survey and the National Household Travel Survey assess how people travel, these surveillance efforts do not assess the surrounding infrastructure which effect travel decisions. Ongoing surveillance of public transportation accessibility infrastructure is necessary for evaluating the impact of public transportation accessibility on health inequities within the population.

However, this project is not without limitations. Scoring was limited to information publicly available on public transportation websites, which may not represent the true experience of how people with disabilities use the system. Additional research is needed to understand how the availability of public transportation accessibility information relates to lived experience (e.g., vigilance, acceptance), health behaviors (e.g., physical activity, diet), individual functioning (e.g., mobility), and social participation (e.g., employment, education). Although website information does not represent the experience of people with disabilities, websites are a primary mode to obtain objective information about public transportation systems and are therefore important for information access. This project was also limited in scope, and only evaluated 26 public transportation agencies in the United States. However, the 26 public transportation agencies evaluated as part of this project received the most federal funding and were among the largest public transportation systems in the United States. Future research is needed to expand data collection to a larger sample of public transportation systems within the United States. Lastly, information gathering about the accessibility of a public transportation system is only one phase of a travel journey chain. Barriers at each phase of the travel journey chain (e.g., information, built environment, public transport) can break the chain and delay, postpone, or cancel a trip.^24^ Additional research is needed to evaluate accessibility at each phase of the travel journey chain to enable equal access for the disability community.

## 5. Conclusions

The TRACT tool was developed to fill a surveillance gap in accessible public transportation systems. Our initial examination found that information about public transportation accessibility varies widely across systems, indicating that the ability to make plans and use public transportation may depend on where riders with disabilities live. This tool can be used to identify and evaluate accessibility information on public transportation system websites, track changes in this information over time, examine relationships with critical health outcomes such as food access, employment, health care access, and participation, and help to establish a more robust national surveillance system of public transportation accessibility.

## Supporting information

Supplement B Data Dictionary

Supplement C Score Generation

Supplement D Syntax

Supplement E Reliability

## Data Availability

All data produced are available online at https://disabilityhealth.jhu.edu/transitdashboard/

## References

1. Varadaraj V, Deal JA, Campanile J, Reed NS, Swenor BK. National prevalence of disability and disability types among adults in the US, 2019. JAMA network open. 2021;4(10):e2130358–e.

2. Americans with Disabilities Act Accessibility Specifications for Transportation Vehicles, 49 C. F. R. Part 38.

3. Transportation Services for Individuals with Disabilities (ADA), 49 C. F. R. Part 37.

4. United States Department of Transportation. ADA Regulations. In: Administration FT, editor. Washington, DC 2012.

5. Bezyak JL, Sabella SA, Gattis RH. Public transportation: an investigation of barriers for people with disabilities. Journal of Disability Policy Studies. 2017;28(1):52–60.

6. Brumbaugh S. Travel patterns of American adults with disabilities. Bur Transp Stat. 2018:1–10.

7. Wolfe MK. Access to Health Care: Perspectives on Transportation as a Social Determinant of Health: The University of North Carolina at Chapel Hill; 2020.

8. McAndrews C, Rosenlieb EG, Troy A, Marshall WE. Transportation and Land Use as Social Determinants of Health: Analysis of Exposure to Traffic in the Denver Metropolitan Region. Mountain-Plains Consortium. 2017;28:5–6.

9. Schwartz N, Buliung R, Wilson K. Disability and food access and insecurity: A scoping review of the literature. Health & place. 2019;57:107–21.

10. Bezyak JL, Sabella S, Hammel J, McDonald K, Jones RA, Barton D. Community participation and public transportation barriers experienced by people with disabilities. Disabil Rehabil. 2020;42(23):3275–83. doi: 10.1080/09638288.2019.1590469. PubMed PMID: 30991852.

11. Syed ST, Gerber BS, Sharp LK. Traveling towards disease: transportation barriers to health care access. Journal of community health. 2013;38(5):976–93.

12. Marrocco A, Krouse HJ. Obstacles to preventive care for individuals with disability: Implications for nurse practitioners. Journal of the American Association of Nurse Practitioners. 2017;29(5):282–93.

13. Statistics BoL. Persons with a disability: Barriers to employment, types of assistance, and other labor related issues. US Department of Labor Washington, DC; 2013.

14. Hammel J, Magasi S, Heinemann A, Gray DB, Stark S, Kisala P, Carlozzi NE, Tulsky D, Garcia SF, Hahn EA. Environmental barriers and supports to everyday participation: a qualitative insider perspective from people with disabilities. Archives of physical medicine and rehabilitation. 2015;96(4):578–88.

15. Remillard ET, Campbell ML, Koon LM, Rogers WA. Transportation challenges for persons aging with mobility disability: Qualitative insights and policy implications. Disability and health journal. 2022;15(1):101209.

16. Office of Civil Rights. ADA Compliance Review Final Reports Washington, DC: Federal Transit Administration [cited 2022 December 7]. Available from: https://www.transit.dot.gov/regulations-and-guidance/civil-rights-ada/ada-compliance-review-final-reports.

17. Steinfeld E, Maisel J. Universal design: Creating inclusive environments: John Wiley & Sons; 2012.

18. Litman T. Evaluating accessibility for transport planning: Victoria Transport Policy Institute Victoria, BC, Canada; 2017.

19. Unsworth C, So MH, Chua J, Gudimetla P, Naweed A. A systematic review of public transport accessibility for people using mobility devices. Disabil Rehabil. 2021;43(16):2253–67. doi: 10.1080/09638288.2019.1697382. PubMed PMID: 31800337.

20. Iwarsson S, Ståhl A. Accessibility, usability and universal design—positioning and definition of concepts describing person-environment relationships. Disability and rehabilitation. 2003;25(2):57–66.

21. Litman T. Evaluating accessibility for transport planning: Victoria Transport Policy Institute Victoria, BC, Canada; 2022.

22. Verseckiene A, Meškauskas V, Batarliene N. Urban public transport accessibility for people with movement disorders: The case study of Vilnius. Procedia Engineering. 2016;134:48–56.

23. National Aging and Disability Transportation Center. NADTC Every Ride Counts Campaign https://www.nadtc.org/wp-content/uploads/Every-Ride-Counts-Webinar-022019.pdf2019 [April 13, 2022]. Available from: https://www.nadtc.org/everyridecounts/.

24. Park J, Chowdhury S. Towards an enabled journey: barriers encountered by public transport riders with disabilities for the whole journey chain. Transport Reviews. 2022;42(2):181–203.

25. Urban Design Working Group. Transit Universal Design Guidelines: Principles and Best Practices for Implementing Universal Design in Transit. 2020 March 3, 2022:[47 p.]. Available from: https://www.apta.com/wp-content/uploads/APTA-SUDS-UD-GL-010-20.pdf.

26. National Aging and Disability Transportation Center. Toolkit for the assessment of bus stop accessibility and safety Washington, D.C. 2014 [cited 2022 Mary 6, 2022]. Available from: https://www.nadtc.org/wp-content/uploads/NADTC-Toolkit-for-the-Assessment-of-Bus-Stop-Accessibility.pdf.

27. 2020 Funding Sources. In: Federal Transit Administration, editor. National Transit Database. Washington, DC 2020.

28. Samuel LJ, Xiao E, Cerilli C, Sweeney F, Campanile J, Milki N, Smith J, Zhu J, Yenokyan G, Gherman A. The development of the Supplemental Nutrition Assistance Program enrollment accessibility (SNAP-Access) score. medRxiv. 2022.

29. WAVE Web Accessibility Evaluation Tools Logan, UT: Institute for Disability Research, Policy, & Practice; 2022 [cited 2022 November 01, 2022]. Available from: https://wave.webaim.org/api/.

30. The WebAIM Million: The 2022 report on the accessibility of the top 1,000,000 home pages. Logan, UT: Institute for Disability Research, Policy, and Practice, 2022.

31. Byrt T, Bishop J, Carlin JB. Bias, prevalence and kappa. Journal of clinical epidemiology. 1993;46(5):423–9.

32. Feinstein AR, Cicchetti DV. High agreement but low kappa: I. The problems of two paradoxes. Journal of clinical epidemiology. 1990;43(6):543–9.

33. Park J, Chowdhury S. Investigating the barriers in a typical journey by public transport users with disabilities. Journal of Transport and Health. 2018;10:361–8. doi: 10.1016/j.jth.2018.05.008.

34. Wayland S, Newland J, Gill-Atkinson L, Vaughan C, Emerson E, Llewellyn G. I had every right to be there: discriminatory acts towards young people with disabilities on public transport. Disability and Society. 2020. doi: 10.1080/09687599.2020.1822784.

35. Frost KL, Bertocci G, Smalley C. Ramps remain a barrier to safe wheelchair user transit bus ingress/egress. Disabil Rehabil Assist Technol. 2020;15(6):629–36. doi: 10.1080/17483107.2019.1604824. PubMed PMID: 32364033.

36. Park J, Chowdhury S. Towards an enabled journey: barriers encountered by public transport riders with disabilities for the whole journey chain. Transport Reviews. 2021. doi: 10.1080/01441647.2021.1955035.

