## Supplement A Figures for "TRansit ACessibility Tool (TRACT): Developing a novel scoring system for public transportation system accessibility"

### Appendix A. Dimension specific horizontal bar charts, the x-axis represents the theoretical range of each dimension and a vertical line is drawn at half of the maximum points for each dimension.

**Figure A.1** TRansit ACcessibility Tool (TRACT) scores for facility accessibility among the top 26 public transportation systems receiving federal funding in the United States.

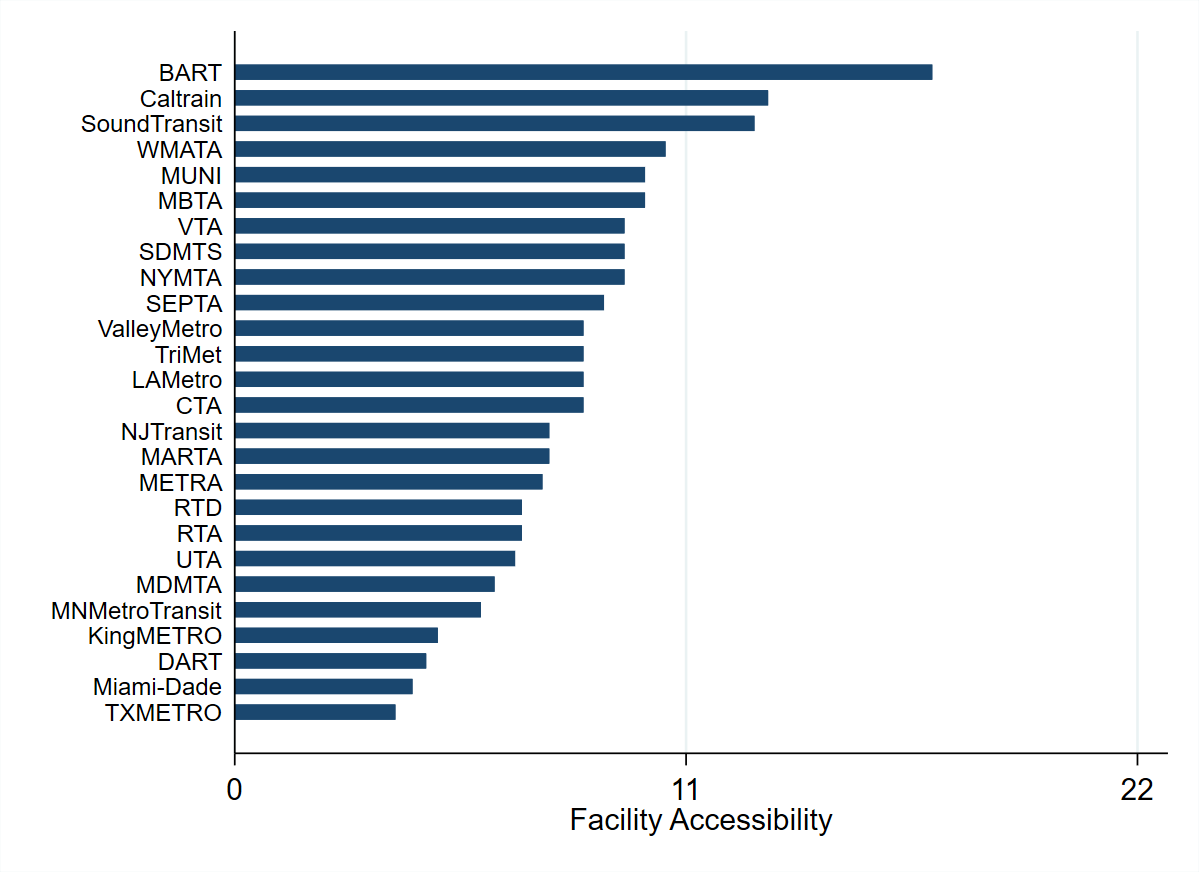

#### Note. In order of appearance: BART = San Francisco Bay Area Rapid Transit District; SoundTransit = Central Puget Sound Regional Transit Authority; WMATA = Washington Metropolitan Area Transit Authority; MUNI = San Francisco Municipal Transportation Agency; MBTA = Massachusetts Bay Transportation Authority; VTA = Santa Clara Valley Transportation Authority; SDMTS = San Diego Metropolitan Transit System; NYMTA = Metropolitan Transit Authority New York City Transit, Metropolitan Transit Authority Long Island Rail Road, Metro-North Commuter Railroad Company; SEPTA = Southeastern Pennsylvania Transportation Authority; ValleyMetro = Valley Metro Rail, Inc.; TriMet = Tri-County Metropolitan Transportation District of Oregon; LAMetro = Los Angeles County Metropolitan Transportation Authority; CTA = Chicago Transit Authority; NJTransit = New Jersey Transit Corporation; MARTA = Metropolitan Atlanta Rapid Transit Authority; METRA = Northeast Illinois Regional Commuter Railroad Corporation; RTD = Denver Regional Transportation District; RTA = The Greater Cleveland Regional Transit Authority; UTA = Utah Transit Authority; MDMTA = Maryland Transit Administration; MNMetroTransit = Minneapolis Minnesota Metro Transit; KingMETRO = King County Department of Metro Transit; DART = Dallas Area Rapid Transit; Miami-Dade = Transportation & Public Work of Miami-Dade TXMETRO = Metropolitan Transit Authority of Harris County, Texas.

#### **Figure A.2** TRansit ACcessibility Tool (TRACT) scores for vehicle accessibility among the top 26 public transportation systems receiving federal funding in the United States.

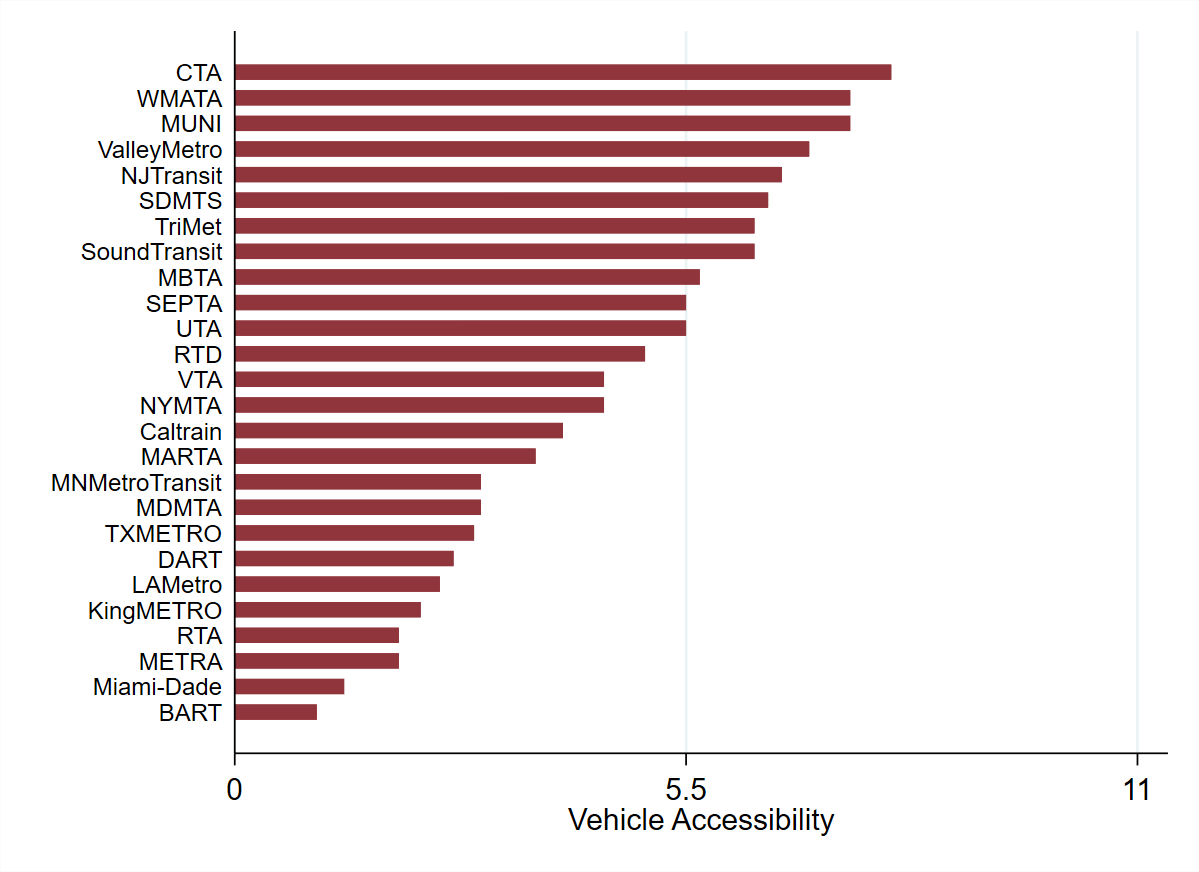

*Note.* In order of appearance: CTA = Chicago Transit Authority; WMATA = Washington Metropolitan Area Transit Authority; MUNI = San Francisco Municipal Transportation Agency; ValleyMetro = Valley Metro Rail, Inc.; NJTransit = New Jersey Transit Corporation; SDMTS = San Diego Metropolitan Transit System; TriMet = Tri-County Metropolitan Transportation District of Oregon; SoundTransit = Central Puget Sound Regional Transit Authority; MBTA = Massachusetts Bay Transportation Authority; SEPTA = Southeastern Pennsylvania Transportation Authority; UTA = Utah Transit Authority; RTD = Denver Regional Transportation District; VTA = Santa Clara Valley Transportation Authority; NYMTA = Metropolitan Transit Authority New York City Transit, Metropolitan Transit Authority Long Island Rail Road, Metro-North Commuter Railroad Company; MARTA = Metropolitan Atlanta Rapid Transit Authority; MNMetroTransit = Minneapolis Minnesota Metro Transit; MDMTA = Maryland Transit Administration; TXMETRO = Metropolitan Transit Authority of Harris County, Texas; DART = Dallas Area Rapid Transit; LAMetro = Los Angeles County Metropolitan Transportation Authority; KingMETRO = King County Department of Metro Transit; RTA = The Greater Cleveland Regional Transit Authority; METRA = Northeast Illinois Regional Commuter Railroad Corporation; Miami-Dade = Transportation & Public Work of Miami-Dade; BART = San Francisco Bay Area Rapid Transit District.

#### **Figure A.3** TRansit ACcessibility Tool (TRACT) scores for inclusive policies among the top 26 public transportation systems receiving federal funding in the United States.

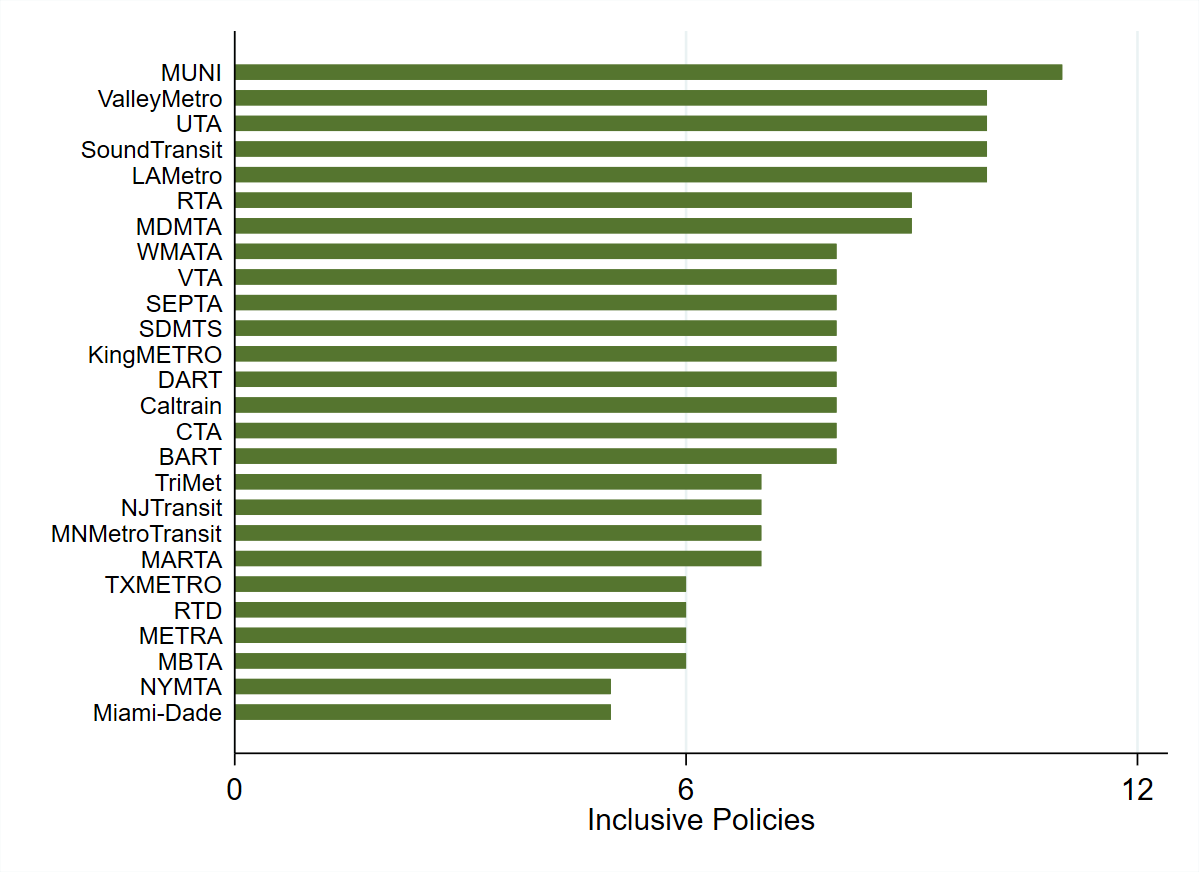

*Note.* In order of appearance: MUNI = San Francisco Municipal Transportation Agency; ValleyMetro = Valley Metro Rail, Inc.; UTA = Utah Transit Authority; SoundTransit = Central Puget Sound Regional Transit Authority; LAMetro = Los Angeles County Metropolitan Transportation Authority; RTA = The Greater Cleveland Regional Transit Authority; MDMTA = Maryland Transit Administration; WMATA = Washington Metropolitan Area Transit Authority; VTA = Santa Clara Valley Transportation Authority; SEPTA = Southeastern Pennsylvania Transportation Authority; SDMTS = San Diego Metropolitan Transit System; KingMETRO = King County Department of Metro Transit; DART = Dallas Area Rapid Transit; CTA = Chicago Transit Authority; BART = San Francisco Bay Area Rapid Transit District; TriMet = Tri-County Metropolitan Transportation District of Oregon; NJTransit = New Jersey Transit Corporation; MNMetroTransit = Minneapolis Minnesota Metro Transit; MARTA = Metropolitan Atlanta Rapid Transit Authority; TXMETRO = Metropolitan Transit Authority of Harris County, Texas; RTD = Denver Regional Transportation District; METRA = Northeast Illinois Regional Commuter Railroad Corporation; MBTA = Massachusetts Bay Transportation Authority; NYMTA = Metropolitan Transit Authority New York City Transit, Metropolitan Transit Authority Long Island Rail Road, Metro-North Commuter Railroad Company; Miami-Dade = Transportation & Public Work of Miami-Dade.

**Figure A.4** TRansit ACcessibility Tool (TRACT) scores for rider accommodations among the top 26 public transportation systems receiving federal funding in the United States.

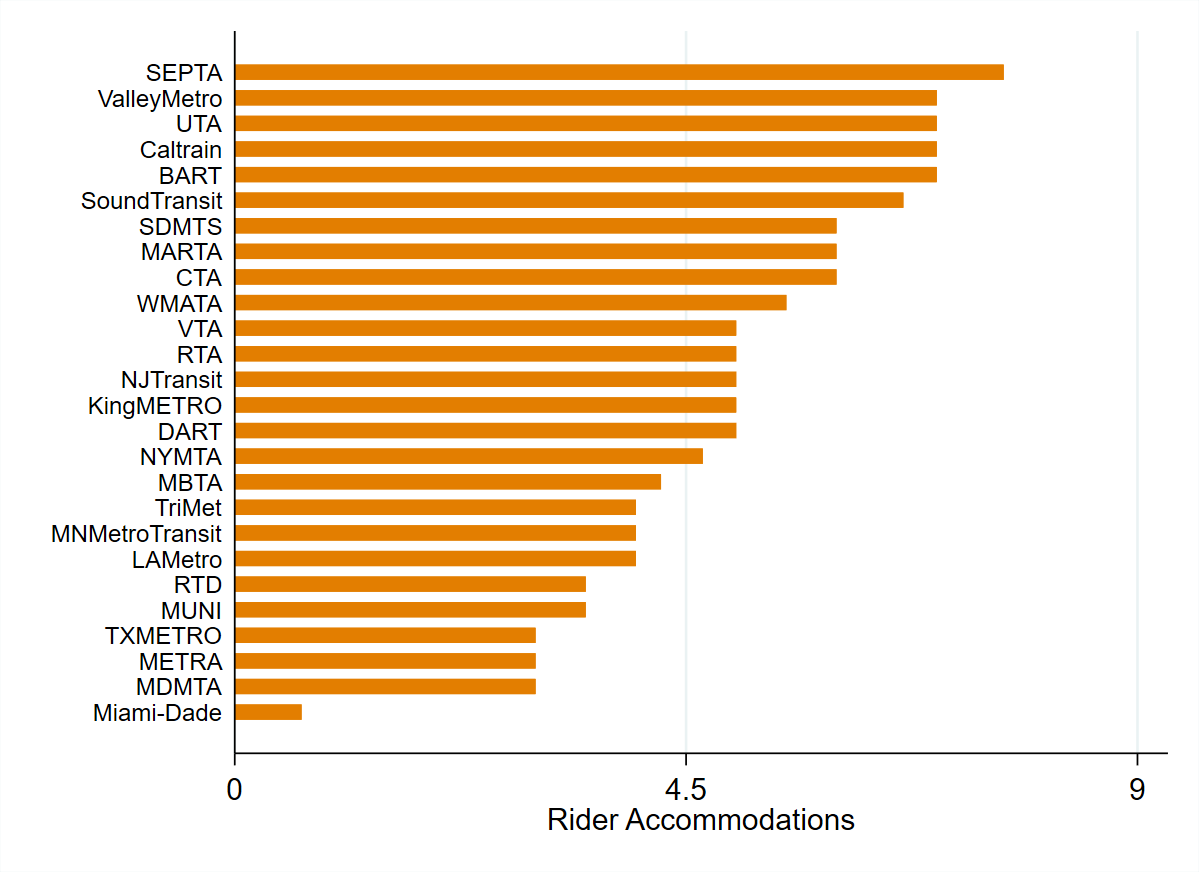

*Note.* In order of appearance: SEPTA = Southeastern Pennsylvania Transportation Authority; ValleyMetro = Valley Metro Rail, Inc.; UTA = Utah Transit Authority; BART = San Francisco Bay Area Rapid Transit District; SoundTransit = Central Puget Sound Regional Transit Authority; SDMTS = San Diego Metropolitan Transit System; MARTA = Metropolitan Atlanta Rapid Transit Authority; CTA = Chicago Transit Authority; WMATA = Washington Metropolitan Area Transit Authority; VTA = Santa Clara Valley Transportation Authority; RTA = The Greater Cleveland Regional Transit Authority; NJTransit = New Jersey Transit Corporation; KingMETRO = King County Department of Metro Transit; DART = Dallas Area Rapid Transit; NYMTA = Metropolitan Transit Authority New York City Transit, Metropolitan Transit Authority Long Island Rail Road, Metro-North Commuter Railroad Company; MBTA = Massachusetts Bay Transportation Authority; TriMet = Tri-County Metropolitan Transportation District of Oregon; MNMetroTransit = Minneapolis Minnesota Metro Transit; LAMetro = Los Angeles County Metropolitan Transportation Authority; RTD = Denver Regional Transportation District; MUNI = San Francisco Municipal Transportation Agency; TXMETRO = Metropolitan Transit Authority of Harris County, Texas; METRA = Northeast Illinois Regional Commuter Railroad Corporation; MDMTA = Maryland Transit Administration; Miami-Dade = Transportation & Public Work of Miami-Dade

**Figure A.5** TRansit ACcessibility Tool (TRACT) scores for paratransit services among the top 26 public transportation systems receiving federal funding in the United States.

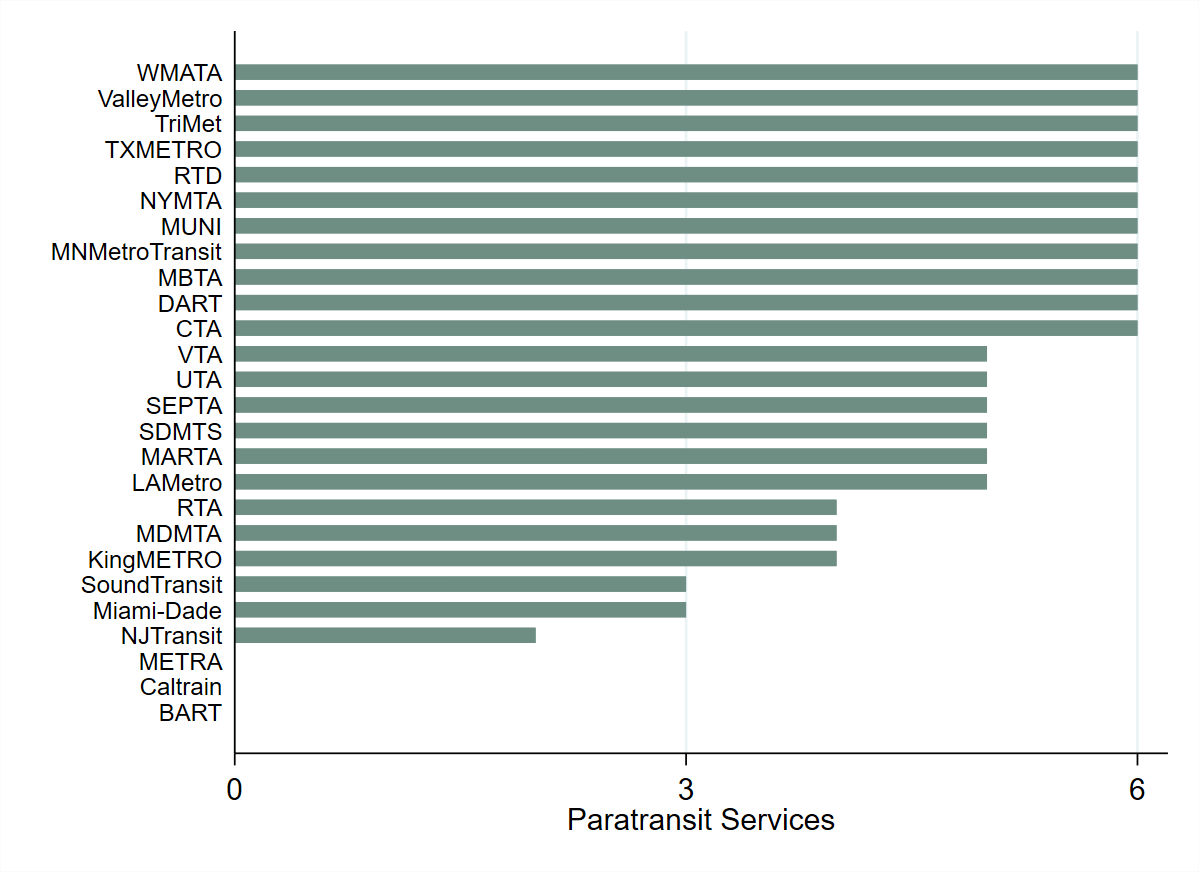

*Note.* In order of appearance: WMATA = Washington Metropolitan Area Transit Authority; ValleyMetro = Valley Metro Rail, Inc.; TriMet = Tri-County Metropolitan Transportation District of Oregon; TXMETRO = Metropolitan Transit Authority of Harris County, Texas; RTD = Denver Regional Transportation District; NYMTA = Metropolitan Transit Authority New York City Transit, Metropolitan Transit Authority Long Island Rail Road, Metro-North Commuter Railroad Company; MUNI = San Francisco Municipal Transportation Agency; MNMetroTransit = Minneapolis Minnesota Metro Transit; MBTA = Massachusetts Bay Transportation Authority; DART = Dallas Area Rapid Transit; CTA = Chicago Transit Authority; VTA = Santa Clara Valley Transportation Authority; UTA = Utah Transit Authority; SEPTA = Southeastern Pennsylvania Transportation Authority; SDMTS = San Diego Metropolitan Transit System; MARTA = Metropolitan Atlanta Rapid Transit Authority; LAMetro = Los Angeles County Metropolitan Transportation Authority; RTA = The Greater Cleveland Regional Transit Authority; MDMTA = Maryland Transit Administration; KingMETRO = King County Department of Metro Transit; SoundTransit = Central Puget Sound Regional Transit Authority; Miami-Dade = Transportation & Public Work of Miami-Dade; NJTransit = New Jersey Transit Corporation; METRA = Northeast Illinois Regional Commuter Railroad Corporation; BART = San Francisco Bay Area Rapid Transit District.

#### **Figure A.6** TRansit ACcessibility Tool (TRACT) scores for website accessibility among the top 26 public transportation systems receiving federal funding in the United States.

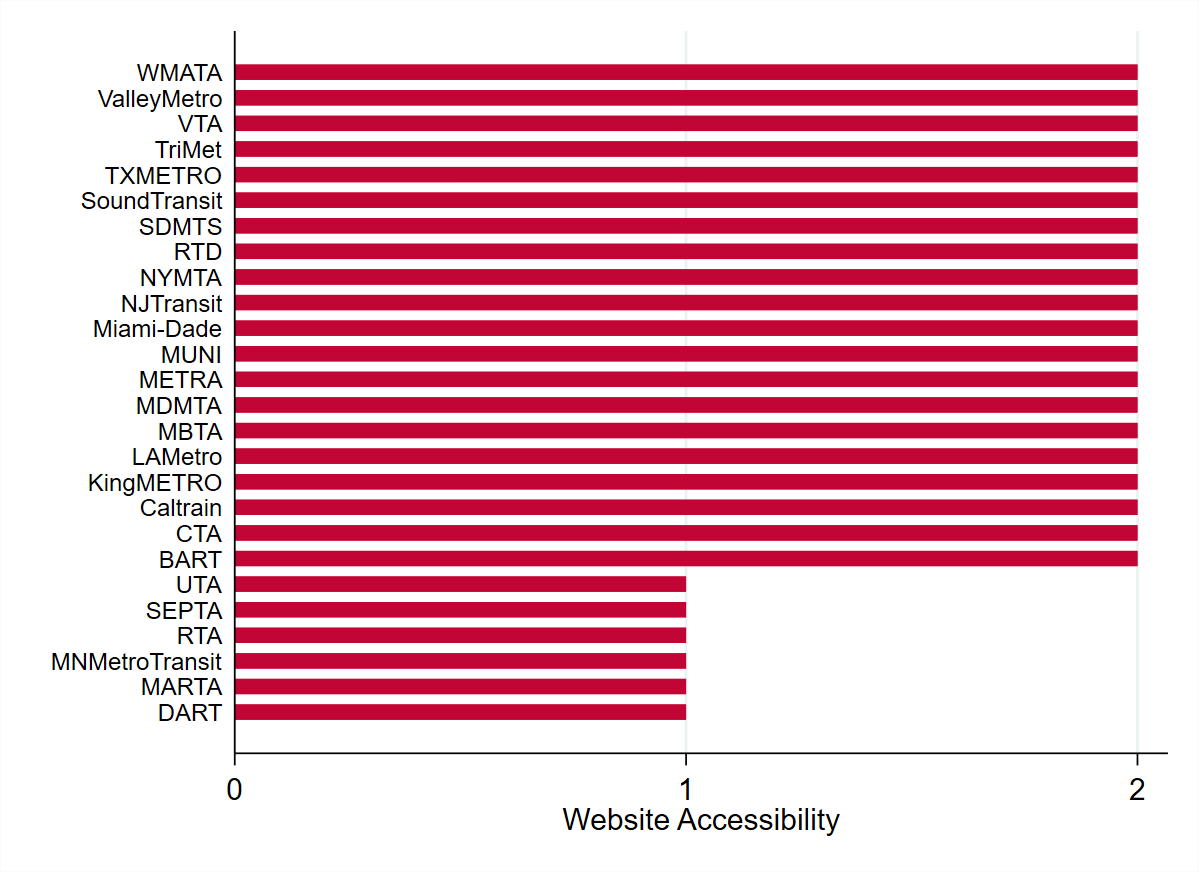

*Note.* In order of appearance: WMATA = Washington Metropolitan Area Transit Authority; ValleyMetro = Valley Metro Rail, Inc.; VTA = Santa Clara Valley Transportation Authority; TriMet = Tri-County Metropolitan Transportation District of Oregon; TXMETRO = Metropolitan Transit Authority of Harris County, Texas; SoundTransit = Central Puget Sound Regional Transit Authority; SDMTS = San Diego Metropolitan Transit System; RTD = Denver Regional Transportation District; NYMTA = Metropolitan Transit Authority New York City Transit, Metropolitan Transit Authority Long Island Rail Road, Metro-North Commuter Railroad Company; NJTransit = New Jersey Transit Corporation; Miami-Dade = Transportation & Public Work of Miami-Dade; MUNI = San Francisco Municipal Transportation Agency; METRA = Northeast Illinois Regional Commuter Railroad Corporation; MDMTA = Maryland Transit Administration; MBTA = Massachusetts Bay Transportation Authority; LAMetro = Los Angeles County Metropolitan Transportation Authority; KingMETRO = King County Department of Metro Transit; CTA = Chicago Transit Authority; BART = San Francisco Bay Area Rapid Transit District; UTA = Utah Transit Authority; SEPTA = Southeastern Pennsylvania Transportation Authority; RTA = The Greater Cleveland Regional Transit Authority; MNMetroTransit = Minneapolis Minnesota Metro Transit; MARTA = Metropolitan Atlanta Rapid Transit Authority; DART = Dallas Area Rapid Transit.
