## Supplement B Data Dictionary for "TRansit ACessibility Tool (TRACT): Developing a novel scoring system for public transportation system accessibility"

### Appendix B. Data entry dictionary with variable names, item content, response options, and coding for all six dimensions and the overall public transportation accessibility information score

#### Table B.1 Facility Accessibility

| Variable name | Item content | Response Options | Coding |
| --- | --- | --- | --- |
| q211_acpr | Which of the following universal design features are listed as available within the BUS FACILITY (e.g. bus stop)? | 0,1 | 0=No Accessible Parking; 1= Accessible Parking |
| q211_acrr | Which of the following universal design features are listed as available within the BUS FACILITY (e.g. bus stop)? | 0,1 | 0=No Accessible Restrooms; 1= Accessible Restrooms |
| q211_altf | Which of the following universal design features are listed as available within the BUS FACILITY (e.g. bus stop)? | 0,1 | 0=No Information in Alternative Formats; 1= Information in Alternative Formats |
| q211_dwar | Which of the following universal design features are listed as available within the BUS FACILITY (e.g. bus stop)? | 0,1 | 0=No Detectable Warnings; 1= Detectable Warnings |
| q211_elev | Which of the following universal design features are listed as available within the BUS FACILITY (e.g. bus stop)? | 0,1 | 0=No Elevator(s); 1= Elevator(s) |
| q211_esca | Which of the following universal design features are listed as available within the BUS FACILITY (e.g. bus stop)? | 0,1 | 0=No Escalator(s); 1= Escalator(s) |
| q211_fltr | Which of the following universal design features are listed as available within the BUS FACILITY (e.g. bus stop)? | 0,1 | 0=No Front Line Staff Training; 1= Front Line Staff Training |
| q211_gate | Which of the following universal design features are listed as available within the BUS FACILITY (e.g. bus stop)? | 0,1 | 0=No Wider Fare Gate(s); 1= Wider Fare Gate(s) |
| q211_hast | Which of the following universal design features are listed as available within the BUS FACILITY (e.g. bus stop)? | 0,1 | 0=No Hearing Assistance System; 1= Hearing Assistance System |
| q211_hlop | Which of the following universal design features are listed as available within the BUS FACILITY (e.g. bus stop)? | 0,1 | 0=No Hearing Loops; 1= Hearing Loops |
| q211_illu | Which of the following universal design features are listed as available within the BUS FACILITY (e.g. bus stop)? | 0,1 | 0=No Illumination of Surfaces; 1= Illumination of Surfaces |
| q211_lvbr | Which of the following universal design features are listed as available within the BUS FACILITY (e.g. bus stop)? | 0,1 | 0=No Level Boarding; 1= Level Boarding |
| q211_nota | Which of the following universal design features are listed as available within the BUS FACILITY (e.g. bus stop)? | 0,1 | 0=At Least One UD Feature; 1= None of the Above |
| q211_ramp | Which of the following universal design features are listed as available within the BUS FACILITY (e.g. bus stop)? | 0,1 | 0=No Ramp(s); 1= Ramp(s) |
| q211_slip | Which of the following universal design features are listed as available within the BUS FACILITY (e.g. bus stop)? | 0,1 | 0=No Slip-Resistant Surfaces; 1= Slip-Resistant Surfaces |
| q211_staa | Which of the following universal design features are listed as available within the BUS FACILITY (e.g. bus stop)? | 0,1 | 0=No Station Audio Announcements; 1= Station Audio Announcements |
| q211_stvi | Which of the following universal design features are listed as available within the BUS FACILITY (e.g. bus stop)? | 0,1 | 0=No Station Visual Information; 1= Station Visual Information |
| q211_tick | Which of the following universal design features are listed as available within the BUS FACILITY (e.g. bus stop)? | 0,1 | 0=No Tickets Available via Kiosk; 1= Tickets Available via Kiosk |
| q211_tico | Which of the following universal design features are listed as available within the BUS FACILITY (e.g. bus stop)? | 0,1 | 0=No Tickets Available Online; 1= Tickets Available Online |
| q211_ticp | Which of the following universal design features are listed as available within the BUS FACILITY (e.g. bus stop)? | 0,1 | 0=No Tickets Available In-Person; 1= Tickets Available In-Person |
| q211_ttpg | Which of the following universal design features are listed as available within the BUS FACILITY (e.g. bus stop)? | 0,1 | 0=No Travel Training Program; 1= Travel Training Program |
| q211_wayf | Which of the following universal design features are listed as available within the BUS FACILITY (e.g. bus stop)? | 0,1 | 0=No Wayfinding Signs; 1= Wayfinding Signs |
| q213_r | Is there information on which BUS FACILITIES (e.g. bus stop) have accessibility features? | 0,1 | 0=No; 1=Yes |
| q221_acpr | Which of the following universal design features are listed as available within the LIGHT RAIL FACILITY (e.g. rail station)? Check all that apply. | 0,1 | 0=No Accessible Parking; 1= Accessible Parking |
| q221_acrr | Which of the following universal design features are listed as available within the LIGHT RAIL FACILITY (e.g. rail station)? Check all that apply. | 0,1 | 0=No Accessible Restrooms; 1= Accessible Restrooms |
| q221_altf | Which of the following universal design features are listed as available within the LIGHT RAIL FACILITY (e.g. rail station)? Check all that apply. | 0,1 | 0=No Information in Alternative Formats; 1= Information in Alternative Formats |
| q221_dwar | Which of the following universal design features are listed as available within the LIGHT RAIL FACILITY (e.g. rail station)? Check all that apply. | 0,1 | 0=No Detectable Warnings; 1= Detectable Warnings |
| q221_elev | Which of the following universal design features are listed as available within the LIGHT RAIL FACILITY (e.g. rail station)? Check all that apply. | 0,1 | 0=No Elevator(s); 1= Elevator(s) |
| q221_esca | Which of the following universal design features are listed as available within the LIGHT RAIL FACILITY (e.g. rail station)? Check all that apply. | 0,1 | 0=No Escalator(s); 1= Escalator(s) |
| q221_fltr | Which of the following universal design features are listed as available within the LIGHT RAIL FACILITY (e.g. rail station)? Check all that apply. | 0,1 | 0=No Front Line Staff Training; 1= Front Line Staff Training |
| q221_gate | Which of the following universal design features are listed as available within the LIGHT RAIL FACILITY (e.g. rail station)? Check all that apply. | 0,1 | 0=No Wider Fare Gate(s); 1= Wider Fare Gate(s) |
| q221_hast | Which of the following universal design features are listed as available within the LIGHT RAIL FACILITY (e.g. rail station)? Check all that apply. | 0,1 | 0=No Hearing Assistance System; 1= Hearing Assistance System |
| q221_hlop | Which of the following universal design features are listed as available within the LIGHT RAIL FACILITY (e.g. rail station)? Check all that apply. | 0,1 | 0=No Hearing Loops; 1= Hearing Loops |
| q221_illu | Which of the following universal design features are listed as available within the LIGHT RAIL FACILITY (e.g. rail station)? Check all that apply. | 0,1 | 0=No Illumination of Surfaces; 1= Illumination of Surfaces |
| q221_lvbr | Which of the following universal design features are listed as available within the LIGHT RAIL FACILITY (e.g. rail station)? Check all that apply. | 0,1 | 0=No Level Boarding; 1= Level Boarding |
| q221_nota | Which of the following universal design features are listed as available within the LIGHT RAIL FACILITY (e.g. rail station)? Check all that apply. | 0,1 | 0=At Least One UD Feature; 1= None of the Above |
| q221_ramp | Which of the following universal design features are listed as available within the LIGHT RAIL FACILITY (e.g. rail station)? Check all that apply. | 0,1 | 0=No Ramp(s); 1= Ramp(s) |
| q221_slip | Which of the following universal design features are listed as available within the LIGHT RAIL FACILITY (e.g. rail station)? Check all that apply. | 0,1 | 0=No Slip-Resistant Surfaces; 1= Slip-Resistant Surfaces |
| q221_staa | Which of the following universal design features are listed as available within the LIGHT RAIL FACILITY (e.g. rail station)? Check all that apply. | 0,1 | 0=No Station Audio Announcements; 1= Station Audio Announcements |
| q221_stvi | Which of the following universal design features are listed as available within the LIGHT RAIL FACILITY (e.g. rail station)? Check all that apply. | 0,1 | 0=No Station Visual Information; 1= Station Visual Information |
| q221_tick | Which of the following universal design features are listed as available within the LIGHT RAIL FACILITY (e.g. rail station)? Check all that apply. | 0,1 | 0=No Tickets Available via Kiosk; 1= Tickets Available via Kiosk |
| q221_tico | Which of the following universal design features are listed as available within the LIGHT RAIL FACILITY (e.g. rail station)? Check all that apply. | 0,1 | 0=No Tickets Available Online; 1= Tickets Available Online |
| q221_ticp | Which of the following universal design features are listed as available within the LIGHT RAIL FACILITY (e.g. rail station)? Check all that apply. | 0,1 | 0=No Tickets Available In-Person; 1= Tickets Available In-Person |
| q221_ttpg | Which of the following universal design features are listed as available within the LIGHT RAIL FACILITY (e.g. rail station)? Check all that apply. | 0,1 | 0=No Travel Training Program; 1= Travel Training Program |
| q221_wayf | Which of the following universal design features are listed as available within the LIGHT RAIL FACILITY (e.g. rail station)? Check all that apply. | 0,1 | 0=No Wayfinding Signs; 1= Wayfinding Signs |
| q223_r | Is there information on which LIGHT RAIL FACILITIES (e.g. train stop) have accessibility features? | 0,1 | 0=No; 1=Yes |
| q231_acpr | Which of the following universal design features are listed as available within the HEAVY RAIL FACILITY (e.g. rail station)? Check all that apply. | 0,1 | 0=No Accessible Parking; 1= Accessible Parking |
| q231_acrr | Which of the following universal design features are listed as available within the HEAVY RAIL FACILITY (e.g. rail station)? Check all that apply. | 0,1 | 0=No Accessible Restrooms; 1= Accessible Restrooms |
| q231_altf | Which of the following universal design features are listed as available within the HEAVY RAIL FACILITY (e.g. rail station)? Check all that apply. | 0,1 | 0=No Information in Alternative Formats; 1= Information in Alternative Formats |
| q231_dwar | Which of the following universal design features are listed as available within the HEAVY RAIL FACILITY (e.g. rail station)? Check all that apply. | 0,1 | 0=No Detectable Warnings; 1= Detectable Warnings |
| q231_elev | Which of the following universal design features are listed as available within the HEAVY RAIL FACILITY (e.g. rail station)? Check all that apply. | 0,1 | 0=No Elevator(s); 1= Elevator(s) |
| q231_esca | Which of the following universal design features are listed as available within the HEAVY RAIL FACILITY (e.g. rail station)? Check all that apply. | 0,1 | 0=No Escalator(s); 1= Escalator(s) |
| q231_fltr | Which of the following universal design features are listed as available within the HEAVY RAIL FACILITY (e.g. rail station)? Check all that apply. | 0,1 | 0=No Front Line Staff Training; 1= Front Line Staff Training |
| q231_gate | Which of the following universal design features are listed as available within the HEAVY RAIL FACILITY (e.g. rail station)? Check all that apply. | 0,1 | 0=No Wider Fare Gate(s); 1= Wider Fare Gate(s) |
| q231_hast | Which of the following universal design features are listed as available within the HEAVY RAIL FACILITY (e.g. rail station)? Check all that apply. | 0,1 | 0=No Hearing Assistance System; 1= Hearing Assistance System |
| q231_hlop | Which of the following universal design features are listed as available within the HEAVY RAIL FACILITY (e.g. rail station)? Check all that apply. | 0,1 | 0=No Hearing Loops; 1= Hearing Loops |
| q231_illu | Which of the following universal design features are listed as available within the HEAVY RAIL FACILITY (e.g. rail station)? Check all that apply. | 0,1 | 0=No Illumination of Surfaces; 1= Illumination of Surfaces |
| q231_lvbr | Which of the following universal design features are listed as available within the HEAVY RAIL FACILITY (e.g. rail station)? Check all that apply. | 0,1 | 0=No Level Boarding; 1= Level Boarding |
| q231_nota | Which of the following universal design features are listed as available within the HEAVY RAIL FACILITY (e.g. rail station)? Check all that apply. | 0,1 | 0=At Least One UD Feature; 1= None of the Above |
| q231_ramp | Which of the following universal design features are listed as available within the HEAVY RAIL FACILITY (e.g. rail station)? Check all that apply. | 0,1 | 0=No Ramp(s); 1= Ramp(s) |
| q231_slip | Which of the following universal design features are listed as available within the HEAVY RAIL FACILITY (e.g. rail station)? Check all that apply. | 0,1 | 0=No Slip-Resistant Surfaces; 1= Slip-Resistant Surfaces |
| q231_staa | Which of the following universal design features are listed as available within the HEAVY RAIL FACILITY (e.g. rail station)? Check all that apply. | 0,1 | 0=No Station Audio Announcements; 1= Station Audio Announcements |
| q231_stvi | Which of the following universal design features are listed as available within the HEAVY RAIL FACILITY (e.g. rail station)? Check all that apply. | 0,1 | 0=No Station Visual Information; 1= Station Visual Information |
| q231_tick | Which of the following universal design features are listed as available within the HEAVY RAIL FACILITY (e.g. rail station)? Check all that apply. | 0,1 | 0=No Tickets Available via Kiosk; 1= Tickets Available via Kiosk |
| q231_tico | Which of the following universal design features are listed as available within the HEAVY RAIL FACILITY (e.g. rail station)? Check all that apply. | 0,1 | 0=No Tickets Available Online; 1= Tickets Available Online |
| q231_ticp | Which of the following universal design features are listed as available within the HEAVY RAIL FACILITY (e.g. rail station)? Check all that apply. | 0,1 | 0=No Tickets Available In-Person; 1= Tickets Available In-Person |
| q231_ttpg | Which of the following universal design features are listed as available within the HEAVY RAIL FACILITY (e.g. rail station)? Check all that apply. | 0,1 | 0=No Travel Training Program; 1= Travel Training Program |
| q231_wayf | Which of the following universal design features are listed as available within the HEAVY RAIL FACILITY (e.g. rail station)? Check all that apply. | 0,1 | 0=No Wayfinding Signs; 1= Wayfinding Signs |
| q233_r | Is there information on which HEAVY RAIL FACILITIES (e.g. train station) have accessibility features? | 0,1 | 0=No; 1=Yes |
| q241_acpr | Which of the following universal design features are listed as available within the Other(1) FACILITY? Check all that apply. | 0,1 | 0=No Accessible Parking; 1= Accessible Parking |
| q241_acrr | Which of the following universal design features are listed as available within the Other(1) FACILITY? Check all that apply. | 0,1 | 0=No Accessible Restrooms; 1= Accessible Restrooms |
| q241_altf | Which of the following universal design features are listed as available within the Other(1) FACILITY? Check all that apply. | 0,1 | 0=No Information in Alternative Formats; 1= Information in Alternative Formats |
| q241_dwar | Which of the following universal design features are listed as available within the Other(1) FACILITY? Check all that apply. | 0,1 | 0=No Detectable Warnings; 1= Detectable Warnings |
| q241_elev | Which of the following universal design features are listed as available within the Other(1) FACILITY? Check all that apply. | 0,1 | 0=No Elevator(s); 1= Elevator(s) |
| q241_esca | Which of the following universal design features are listed as available within the Other(1) FACILITY? Check all that apply. | 0,1 | 0=No Escalator(s); 1= Escalator(s) |
| q241_fltr | Which of the following universal design features are listed as available within the Other(1) FACILITY? Check all that apply. | 0,1 | 0=No Front Line Staff Training; 1= Front Line Staff Training |
| q241_gate | Which of the following universal design features are listed as available within the Other(1) FACILITY? Check all that apply. | 0,1 | 0=No Wider Fare Gate(s); 1= Wider Fare Gate(s) |
| q241_hast | Which of the following universal design features are listed as available within the Other(1) FACILITY? Check all that apply. | 0,1 | 0=No Hearing Assistance System; 1= Hearing Assistance System |
| q241_hlop | Which of the following universal design features are listed as available within the Other(1) FACILITY? Check all that apply. | 0,1 | 0=No Hearing Loops; 1= Hearing Loops |
| q241_illu | Which of the following universal design features are listed as available within the Other(1) FACILITY? Check all that apply. | 0,1 | 0=No Illumination of Surfaces; 1= Illumination of Surfaces |
| q241_lvbr | Which of the following universal design features are listed as available within the Other(1) FACILITY? Check all that apply. | 0,1 | 0=No Level Boarding; 1= Level Boarding |
| q241_nota | Which of the following universal design features are listed as available within the Other(1) FACILITY? Check all that apply. | 0,1 | 0=At Least One UD Feature; 1= None of the Above |
| q241_ramp | Which of the following universal design features are listed as available within the Other(1) FACILITY? Check all that apply. | 0,1 | 0=No Ramp(s); 1= Ramp(s) |
| q241_slip | Which of the following universal design features are listed as available within the Other(1) FACILITY? Check all that apply. | 0,1 | 0=No Slip-Resistant Surfaces; 1= Slip-Resistant Surfaces |
| q241_staa | Which of the following universal design features are listed as available within the Other(1) FACILITY? Check all that apply. | 0,1 | 0=No Station Audio Announcements; 1= Station Audio Announcements |
| q241_stvi | Which of the following universal design features are listed as available within the Other(1) FACILITY? Check all that apply. | 0,1 | 0=No Station Visual Information; 1= Station Visual Information |
| q241_tick | Which of the following universal design features are listed as available within the Other(1) FACILITY? Check all that apply. | 0,1 | 0=No Tickets Available via Kiosk; 1= Tickets Available via Kiosk |
| q241_tico | Which of the following universal design features are listed as available within the Other(1) FACILITY? Check all that apply. | 0,1 | 0=No Tickets Available Online; 1= Tickets Available Online |
| q241_ticp | Which of the following universal design features are listed as available within the Other(1) FACILITY? Check all that apply. | 0,1 | 0=No Tickets Available In-Person; 1= Tickets Available In-Person |
| q241_ttpg | Which of the following universal design features are listed as available within the Other(1) FACILITY? Check all that apply. | 0,1 | 0=No Travel Training Program; 1= Travel Training Program |
| q241_wayf | Which of the following universal design features are listed as available within the Other(1) FACILITY? Check all that apply. | 0,1 | 0=No Wayfinding Signs; 1= Wayfinding Signs |
| q243_r | Is there information on which Other(1) FACILITIES (e.g. stop, station, etc.) have accessibility features? | 0,1 | 0=No; 1=Yes |
| q251_acpr | Which of the following universal design features are listed as available within the Other(2) FACILITY? Check all that apply. | 0,1 | 0=No Accessible Parking; 1= Accessible Parking |
| q251_acrr | Which of the following universal design features are listed as available within the Other(2) FACILITY? Check all that apply. | 0,1 | 0=No Accessible Restrooms; 1= Accessible Restrooms |
| q251_altf | Which of the following universal design features are listed as available within the Other(2) FACILITY? Check all that apply. | 0,1 | 0=No Information in Alternative Formats; 1= Information in Alternative Formats |
| q251_dwar | Which of the following universal design features are listed as available within the Other(2) FACILITY? Check all that apply. | 0,1 | 0=No Detectable Warnings; 1= Detectable Warnings |
| q251_elev | Which of the following universal design features are listed as available within the Other(2) FACILITY? Check all that apply. | 0,1 | 0=No Elevator(s); 1= Elevator(s) |
| q251_esca | Which of the following universal design features are listed as available within the Other(2) FACILITY? Check all that apply. | 0,1 | 0=No Escalator(s); 1= Escalator(s) |
| q251_fltr | Which of the following universal design features are listed as available within the Other(2) FACILITY? Check all that apply. | 0,1 | 0=No Front Line Staff Training; 1= Front Line Staff Training |
| q251_gate | Which of the following universal design features are listed as available within the Other(2) FACILITY? Check all that apply. | 0,1 | 0=No Wider Fare Gate(s); 1= Wider Fare Gate(s) |
| q251_hast | Which of the following universal design features are listed as available within the Other(2) FACILITY? Check all that apply. | 0,1 | 0=No Hearing Assistance System; 1= Hearing Assistance System |
| q251_hlop | Which of the following universal design features are listed as available within the Other(2) FACILITY? Check all that apply. | 0,1 | 0=No Hearing Loops; 1= Hearing Loops |
| q251_illu | Which of the following universal design features are listed as available within the Other(2) FACILITY? Check all that apply. | 0,1 | 0=No Illumination of Surfaces; 1= Illumination of Surfaces |
| q251_lvbr | Which of the following universal design features are listed as available within the Other(2) FACILITY? Check all that apply. | 0,1 | 0=No Level Boarding; 1= Level Boarding |
| q251_nota | Which of the following universal design features are listed as available within the Other(2) FACILITY? Check all that apply. | 0,1 | 0=At Least One UD Feature; 1= None of the Above |
| q251_ramp | Which of the following universal design features are listed as available within the Other(2) FACILITY? Check all that apply. | 0,1 | 0=No Ramp(s); 1= Ramp(s) |
| q251_slip | Which of the following universal design features are listed as available within the Other(2) FACILITY? Check all that apply. | 0,1 | 0=No Slip-Resistant Surfaces; 1= Slip-Resistant Surfaces |
| q251_staa | Which of the following universal design features are listed as available within the Other(2) FACILITY? Check all that apply. | 0,1 | 0=No Station Audio Announcements; 1= Station Audio Announcements |
| q251_stvi | Which of the following universal design features are listed as available within the Other(2) FACILITY? Check all that apply. | 0,1 | 0=No Station Visual Information; 1= Station Visual Information |
| q251_tick | Which of the following universal design features are listed as available within the Other(2) FACILITY? Check all that apply. | 0,1 | 0=No Tickets Available via Kiosk; 1= Tickets Available via Kiosk |
| q251_tico | Which of the following universal design features are listed as available within the Other(2) FACILITY? Check all that apply. | 0,1 | 0=No Tickets Available Online; 1= Tickets Available Online |
| q251_ticp | Which of the following universal design features are listed as available within the Other(2) FACILITY? Check all that apply. | 0,1 | 0=No Tickets Available In-Person; 1= Tickets Available In-Person |
| q251_ttpg | Which of the following universal design features are listed as available within the Other(2) FACILITY? Check all that apply. | 0,1 | 0=No Travel Training Program; 1= Travel Training Program |
| q251_wayf | Which of the following universal design features are listed as available within the Other(2) FACILITY? Check all that apply. | 0,1 | 0=No Wayfinding Signs; 1= Wayfinding Signs |
| q253_r | Is there information on which Other(2) FACILITIES (e.g. stop, station, etc.) have accessibility features? | 0,1 | 0=No; 1=Yes |
| q261_acpr | Which of the following universal design features are listed as available within the Other(3) FACILITY? Check all that apply. | 0,1 | 0=No Accessible Parking; 1= Accessible Parking |
| q261_acrr | Which of the following universal design features are listed as available within the Other(3) FACILITY? Check all that apply. | 0,1 | 0=No Accessible Restrooms; 1= Accessible Restrooms |
| q261_altf | Which of the following universal design features are listed as available within the Other(3) FACILITY? Check all that apply. | 0,1 | 0=No Information in Alternative Formats; 1= Information in Alternative Formats |
| q261_dwar | Which of the following universal design features are listed as available within the Other(3) FACILITY? Check all that apply. | 0,1 | 0=No Detectable Warnings; 1= Detectable Warnings |
| q261_elev | Which of the following universal design features are listed as available within the Other(3) FACILITY? Check all that apply. | 0,1 | 0=No Elevator(s); 1= Elevator(s) |
| q261_esca | Which of the following universal design features are listed as available within the Other(3) FACILITY? Check all that apply. | 0,1 | 0=No Escalator(s); 1= Escalator(s) |
| q261_fltr | Which of the following universal design features are listed as available within the Other(3) FACILITY? Check all that apply. | 0,1 | 0=No Front Line Staff Training; 1= Front Line Staff Training |
| q261_gate | Which of the following universal design features are listed as available within the Other(3) FACILITY? Check all that apply. | 0,1 | 0=No Wider Fare Gate(s); 1= Wider Fare Gate(s) |
| q261_hast | Which of the following universal design features are listed as available within the Other(3) FACILITY? Check all that apply. | 0,1 | 0=No Hearing Assistance System; 1= Hearing Assistance System |
| q261_hlop | Which of the following universal design features are listed as available within the Other(3) FACILITY? Check all that apply. | 0,1 | 0=No Hearing Loops; 1= Hearing Loops |
| q261_illu | Which of the following universal design features are listed as available within the Other(3) FACILITY? Check all that apply. | 0,1 | 0=No Illumination of Surfaces; 1= Illumination of Surfaces |
| q261_lvbr | Which of the following universal design features are listed as available within the Other(3) FACILITY? Check all that apply. | 0,1 | 0=No Level Boarding; 1= Level Boarding |
| q261_nota | Which of the following universal design features are listed as available within the Other(3) FACILITY? Check all that apply. | 0,1 | 0=At Least One UD Feature; 1= None of the Above |
| q261_ramp | Which of the following universal design features are listed as available within the Other(3) FACILITY? Check all that apply. | 0,1 | 0=No Ramp(s); 1= Ramp(s) |
| q261_slip | Which of the following universal design features are listed as available within the Other(3) FACILITY? Check all that apply. | 0,1 | 0=No Slip-Resistant Surfaces; 1= Slip-Resistant Surfaces |
| q261_staa | Which of the following universal design features are listed as available within the Other(3) FACILITY? Check all that apply. | 0,1 | 0=No Station Audio Announcements; 1= Station Audio Announcements |
| q261_stvi | Which of the following universal design features are listed as available within the Other(3) FACILITY? Check all that apply. | 0,1 | 0=No Station Visual Information; 1= Station Visual Information |
| q261_tick | Which of the following universal design features are listed as available within the Other(3) FACILITY? Check all that apply. | 0,1 | 0=No Tickets Available via Kiosk; 1= Tickets Available via Kiosk |
| q261_tico | Which of the following universal design features are listed as available within the Other(3) FACILITY? Check all that apply. | 0,1 | 0=No Tickets Available Online; 1= Tickets Available Online |
| q261_ticp | Which of the following universal design features are listed as available within the Other(3) FACILITY? Check all that apply. | 0,1 | 0=No Tickets Available In-Person; 1= Tickets Available In-Person |
| q261_ttpg | Which of the following universal design features are listed as available within the Other(3) FACILITY? Check all that apply. | 0,1 | 0=No Travel Training Program; 1= Travel Training Program |
| q261_wayf | Which of the following universal design features are listed as available within the Other(3) FACILITY? Check all that apply. | 0,1 | 0=No Wayfinding Signs; 1= Wayfinding Signs |
| q263_r | Is there information on which Other(3) FACILITIES (e.g. stop, station, etc.) have accessibility features? | 0,1 | 0=No; 1=Yes |

#### Table B.2 Vehicle Accessibility

| Variable name | Item content | Response Options | Coding |
| --- | --- | --- | --- |
| q214_dwar | Which of the following accessibility features are listed as available on or within the BUS VEHICLES? Check all that apply. | 0,1 | 0=No Detectable Warnings; 1= Detectable Warnings |
| q214_exin | Which of the following accessibility features are listed as available on or within the BUS VEHICLES? Check all that apply. | 0,1 | 0=No External Bus Information; 1= External Bus Information |
| q214_gapr | Which of the following accessibility features are listed as available on or within the BUS VEHICLES? Check all that apply. | 0,1 | 0=No Gap Reducers; 1= Gap Reducers |
| q214_lben | Which of the following accessibility features are listed as available on or within the BUS VEHICLES? Check all that apply. | 0,1 | 0=No Lower Body Entrance; 1= Lower Body Entrance |
| q214_lift | Which of the following accessibility features are listed as available on or within the BUS VEHICLES? Check all that apply. | 0,1 | 0=No Lift(s); 1= Lift(s) |
| q214_nota | Which of the following accessibility features are listed as available on or within the BUS VEHICLES? Check all that apply. | 0,1 | 0=At Least One UD Feature; 1= None of the Above |
| q214_oban | Which of the following accessibility features are listed as available on or within the BUS VEHICLES? Check all that apply. | 0,1 | 0=No On-Board Audio Announcements; 1= On-Board Audio Announcements |
| q214_obvi | Which of the following accessibility features are listed as available on or within the BUS VEHICLES? Check all that apply. | 0,1 | 0=No On-Board Visual Information; 1= On-Board Visual Information |
| q214_oth | Which of the following accessibility features are listed as available on or within the BUS VEHICLES? Check all that apply. | 0,1 | 0=No Other Universal Design Features; 1= Other Universal Design Features |
| q214_ramp | Which of the following accessibility features are listed as available on or within the BUS VEHICLES? Check all that apply. | 0,1 | 0=No Ramp(s); 1= Ramp(s) |
| q214_secu | Which of the following accessibility features are listed as available on or within the BUS VEHICLES? Check all that apply. | 0,1 | 0=No Securement Devices; 1= Securement Devices |
| q214_vsrs | Which of the following accessibility features are listed as available on or within the BUS VEHICLES? Check all that apply. | 0,1 | 0=No Variation in Stop Request Signal; 1= Variation in Stop Request Signal |
| q216_r | Is there information on which BUS VEHICLES (e.g. fleet, line, etc.) have accessibility features? | 0,1 | 0=No; 1=Yes |
| q224_dwar | Which of the following accessibility features are listed as available on or within the LIGHT RAIL VEHICLES? Check all that apply. | 0,1 | 0=No Detectable Warnings; 1= Detectable Warnings |
| q224_exin | Which of the following accessibility features are listed as available on or within the LIGHT RAIL VEHICLES? Check all that apply. | 0,1 | 0=No External Bus Information; 1= External Bus Information |
| q224_gapr | Which of the following accessibility features are listed as available on or within the LIGHT RAIL VEHICLES? Check all that apply. | 0,1 | 0=No Gap Reducers; 1= Gap Reducers |
| q224_lben | Which of the following accessibility features are listed as available on or within the LIGHT RAIL VEHICLES? Check all that apply. | 0,1 | 0=No Lower Body Entrance; 1= Lower Body Entrance |
| q224_lift | Which of the following accessibility features are listed as available on or within the LIGHT RAIL VEHICLES? Check all that apply. | 0,1 | 0=No Lift(s); 1= Lift(s) |
| q224_nota | Which of the following accessibility features are listed as available on or within the LIGHT RAIL VEHICLES? Check all that apply. | 0,1 | 0=At Least One UD Feature; 1= None of the Above |
| q224_oban | Which of the following accessibility features are listed as available on or within the LIGHT RAIL VEHICLES? Check all that apply. | 0,1 | 0=No On-Board Audio Announcements; 1= On-Board Audio Announcements |
| q224_obvi | Which of the following accessibility features are listed as available on or within the LIGHT RAIL VEHICLES? Check all that apply. | 0,1 | 0=No On-Board Visual Information; 1= On-Board Visual Information |
| q224_oth | Which of the following accessibility features are listed as available on or within the LIGHT RAIL VEHICLES? Check all that apply. | 0,1 | 0=No Other Universal Design Features; 1= Other Universal Design Features |
| q224_ramp | Which of the following accessibility features are listed as available on or within the LIGHT RAIL VEHICLES? Check all that apply. | 0,1 | 0=No Ramp(s); 1= Ramp(s) |
| q224_secu | Which of the following accessibility features are listed as available on or within the LIGHT RAIL VEHICLES? Check all that apply. | 0,1 | 0=No Securement Devices; 1= Securement Devices |
| q224_vsrs | Which of the following accessibility features are listed as available on or within the LIGHT RAIL VEHICLES? Check all that apply. | 0,1 | 0=No Variation in Stop Request Signal; 1= Variation in Stop Request Signal |
| q226_r | Is there information on which LIGHT RAIL VEHICLES (e.g. fleet, line, etc.) have accessibility features? | 0,1 | 0=No; 1=Yes |
| q234_dwar | Which of the following accessibility features are listed as available on or within the HEAVY RAIL VEHICLES? Check all that apply. | 0,1 | 0=No Detectable Warnings; 1= Detectable Warnings |
| q234_exin | Which of the following accessibility features are listed as available on or within the HEAVY RAIL VEHICLES? Check all that apply. | 0,1 | 0=No External Bus Information; 1= External Bus Information |
| q234_gapr | Which of the following accessibility features are listed as available on or within the HEAVY RAIL VEHICLES? Check all that apply. | 0,1 | 0=No Gap Reducers; 1= Gap Reducers |
| q234_lben | Which of the following accessibility features are listed as available on or within the HEAVY RAIL VEHICLES? Check all that apply. | 0,1 | 0=No Lower Body Entrance; 1= Lower Body Entrance |
| q234_lift | Which of the following accessibility features are listed as available on or within the HEAVY RAIL VEHICLES? Check all that apply. | 0,1 | 0=No Lift(s); 1= Lift(s) |
| q234_nota | Which of the following accessibility features are listed as available on or within the HEAVY RAIL VEHICLES? Check all that apply. | 0,1 | 0=At Least One UD Feature; 1= None of the Above |
| q234_oban | Which of the following accessibility features are listed as available on or within the HEAVY RAIL VEHICLES? Check all that apply. | 0,1 | 0=No On-Board Audio Announcements; 1= On-Board Audio Announcements |
| q234_obvi | Which of the following accessibility features are listed as available on or within the HEAVY RAIL VEHICLES? Check all that apply. | 0,1 | 0=No On-Board Visual Information; 1= On-Board Visual Information |
| q234_oth | Which of the following accessibility features are listed as available on or within the HEAVY RAIL VEHICLES? Check all that apply. | 0,1 | 0=No Other Universal Design Features; 1= Other Universal Design Features |
| q234_ramp | Which of the following accessibility features are listed as available on or within the HEAVY RAIL VEHICLES? Check all that apply. | 0,1 | 0=No Ramp(s); 1= Ramp(s) |
| q234_secu | Which of the following accessibility features are listed as available on or within the HEAVY RAIL VEHICLES? Check all that apply. | 0,1 | 0=No Securement Devices; 1= Securement Devices |
| q234_vsrs | Which of the following accessibility features are listed as available on or within the HEAVY RAIL VEHICLES? Check all that apply. | 0,1 | 0=No Variation in Stop Request Signal; 1= Variation in Stop Request Signal |
| q236_r | Is there information on which HEAVY RAIL VEHICLES (e.g. fleet, line, etc.) have accessibility features? | 0,1 | 0=No; 1=Yes |
| q244_dwar | Which of the following accessibility features are listed as available on or within the Other(1) VEHICLES? Check all that apply. | 0,1 | 0=No Detectable Warnings; 1= Detectable Warnings |
| q244_exin | Which of the following accessibility features are listed as available on or within the Other(1) VEHICLES? Check all that apply. | 0,1 | 0=No External Bus Information; 1= External Bus Information |
| q244_gapr | Which of the following accessibility features are listed as available on or within the Other(1) VEHICLES? Check all that apply. | 0,1 | 0=No Gap Reducers; 1= Gap Reducers |
| q244_lben | Which of the following accessibility features are listed as available on or within the Other(1) VEHICLES? Check all that apply. | 0,1 | 0=No Lower Body Entrance; 1= Lower Body Entrance |
| q244_lift | Which of the following accessibility features are listed as available on or within the Other(1) VEHICLES? Check all that apply. | 0,1 | 0=No Lift(s); 1= Lift(s) |
| q244_nota | Which of the following accessibility features are listed as available on or within the Other(1) VEHICLES? Check all that apply. | 0,1 | 0=At Least One UD Feature; 1= None of the Above |
| q244_oban | Which of the following accessibility features are listed as available on or within the Other(1) VEHICLES? Check all that apply. | 0,1 | 0=No On-Board Audio Announcements; 1= On-Board Audio Announcements |
| q244_obvi | Which of the following accessibility features are listed as available on or within the Other(1) VEHICLES? Check all that apply. | 0,1 | 0=No On-Board Visual Information; 1= On-Board Visual Information |
| q244_oth | Which of the following accessibility features are listed as available on or within the Other(1) VEHICLES? Check all that apply. | 0,1 | 0=No Other Universal Design Features; 1= Other Universal Design Features |
| q244_ramp | Which of the following accessibility features are listed as available on or within the Other(1) VEHICLES? Check all that apply. | 0,1 | 0=No Ramp(s); 1= Ramp(s) |
| q244_secu | Which of the following accessibility features are listed as available on or within the Other(1) VEHICLES? Check all that apply. | 0,1 | 0=No Securement Devices; 1= Securement Devices |
| q244_vsrs | Which of the following accessibility features are listed as available on or within the Other(1) VEHICLES? Check all that apply. | 0,1 | 0=No Variation in Stop Request Signal; 1= Variation in Stop Request Signal |
| q246_r | Is there information on which Other(1) VEHICLES (e.g. fleet, line, etc.) have accessibility features? | 0,1 | 0=No; 1=Yes |
| q254_dwar | Which of the following accessibility features are listed as available on or within the Other(2) VEHICLES? Check all that apply. | 0,1 | 0=No Detectable Warnings; 1= Detectable Warnings |
| q254_exin | Which of the following accessibility features are listed as available on or within the Other(2) VEHICLES? Check all that apply. | 0,1 | 0=No External Bus Information; 1= External Bus Information |
| q254_gapr | Which of the following accessibility features are listed as available on or within the Other(2) VEHICLES? Check all that apply. | 0,1 | 0=No Gap Reducers; 1= Gap Reducers |
| q254_lben | Which of the following accessibility features are listed as available on or within the Other(2) VEHICLES? Check all that apply. | 0,1 | 0=No Lower Body Entrance; 1= Lower Body Entrance |
| q254_lift | Which of the following accessibility features are listed as available on or within the Other(2) VEHICLES? Check all that apply. | 0,1 | 0=No Lift(s); 1= Lift(s) |
| q254_nota | Which of the following accessibility features are listed as available on or within the Other(2) VEHICLES? Check all that apply. | 0,1 | 0=At Least One UD Feature; 1= None of the Above |
| q254_oban | Which of the following accessibility features are listed as available on or within the Other(2) VEHICLES? Check all that apply. | 0,1 | 0=No On-Board Audio Announcements; 1= On-Board Audio Announcements |
| q254_obvi | Which of the following accessibility features are listed as available on or within the Other(2) VEHICLES? Check all that apply. | 0,1 | 0=No On-Board Visual Information; 1= On-Board Visual Information |
| q254_oth | Which of the following accessibility features are listed as available on or within the Other(2) VEHICLES? Check all that apply. | 0,1 | 0=No Other Universal Design Features; 1= Other Universal Design Features |
| q254_ramp | Which of the following accessibility features are listed as available on or within the Other(2) VEHICLES? Check all that apply. | 0,1 | 0=No Ramp(s); 1= Ramp(s) |
| q254_secu | Which of the following accessibility features are listed as available on or within the Other(2) VEHICLES? Check all that apply. | 0,1 | 0=No Securement Devices; 1= Securement Devices |
| q254_vsrs | Which of the following accessibility features are listed as available on or within the Other(2) VEHICLES? Check all that apply. | 0,1 | 0=No Variation in Stop Request Signal; 1= Variation in Stop Request Signal |
| q256_r | Is there information on which Other(2) VEHICLES (e.g. fleet, line, etc.) have accessibility features? | 0,1 | 0=No; 1=Yes |
| q264_dwar | Which of the following accessibility features are listed as available on or within the Other(3) VEHICLES? Check all that apply. | 0,1 | 0=No Detectable Warnings; 1= Detectable Warnings |
| q264_exin | Which of the following accessibility features are listed as available on or within the Other(3) VEHICLES? Check all that apply. | 0,1 | 0=No External Bus Information; 1= External Bus Information |
| q264_gapr | Which of the following accessibility features are listed as available on or within the Other(3) VEHICLES? Check all that apply. | 0,1 | 0=No Gap Reducers; 1= Gap Reducers |
| q264_lben | Which of the following accessibility features are listed as available on or within the Other(3) VEHICLES? Check all that apply. | 0,1 | 0=No Lower Body Entrance; 1= Lower Body Entrance |
| q264_lift | Which of the following accessibility features are listed as available on or within the Other(3) VEHICLES? Check all that apply. | 0,1 | 0=No Lift(s); 1= Lift(s) |
| q264_nota | Which of the following accessibility features are listed as available on or within the Other(3) VEHICLES? Check all that apply. | 0,1 | 0=At Least One UD Feature; 1= None of the Above |
| q264_oban | Which of the following accessibility features are listed as available on or within the Other(3) VEHICLES? Check all that apply. | 0,1 | 0=No On-Board Audio Announcements; 1= On-Board Audio Announcements |
| q264_obvi | Which of the following accessibility features are listed as available on or within the Other(3) VEHICLES? Check all that apply. | 0,1 | 0=No On-Board Visual Information; 1= On-Board Visual Information |
| q264_oth | Which of the following accessibility features are listed as available on or within the Other(3) VEHICLES? Check all that apply. | 0,1 | 0=No Other Universal Design Features; 1= Other Universal Design Features |
| q264_ramp | Which of the following accessibility features are listed as available on or within the Other(3) VEHICLES? Check all that apply. | 0,1 | 0=No Ramp(s); 1= Ramp(s) |
| q264_secu | Which of the following accessibility features are listed as available on or within the Other(3) VEHICLES? Check all that apply. | 0,1 | 0=No Securement Devices; 1= Securement Devices |
| q264_vsrs | Which of the following accessibility features are listed as available on or within the Other(3) VEHICLES? Check all that apply. | 0,1 | 0=No Variation in Stop Request Signal; 1= Variation in Stop Request Signal |
| q266_r | Is there information on which Other(3) VEHICLES (e.g. fleet, line, etc.) have accessibility features? | 0,1 | 0=No; 1=Yes |

#### Table B.3 Inclusive Policies

| Variable name | Item content | Response Options | Coding |
| --- | --- | --- | --- |
| q19_r | Does the public transit landing page have information describing accessibility policy (e.g. ADA compliance)? | 0,1 | 0=No; 1=Yes |
| q36_r | Does the website provide information about how to submit a complaint? | 0,1 | 0=No; 1=Yes |
| q37_r | Is the designated employee responsible for prompt and equitable resolution of complaints listed? | 0,1 | 0=No; 1=Yes |
| q38_emai | What method is used to submit a complaint? Check all that apply. | 0,1 | 0=No Email; 1=Email |
| q38_iper | What method is used to submit a complaint? Check all that apply. | 0,1 | 0=No In-Person; 1=In-Person |
| q38_mail | What method is used to submit a complaint? Check all that apply. | 0,1 | 0=No Mail; 1=Mail |
| q38_noinfo | What method is used to submit a complaint? Check all that apply. | 0,1 | 0=Information Available; 1=No Information Available |
| q38_oth | What method is used to submit a complaint? Check all that apply. | 0,1 | 0=No Other; 1=Other |
| q38_tele | What method is used to submit a complaint? Check all that apply. | 0,1 | 0=No Telephone; 1=Telephone |
| q38_webf | What method is used to submit a complaint? Check all that apply. | 0,1 | 0=No Web-based Form; 1=Web-based Form |
| q310_r | Telephone is used to submit a complaint, is text telephone (TTY) available? | 0,1 | 0=No; 1=Yes |
| q312_r | Is the name and contact information of the ADA coordinator listed in the public transportation website? | 0,1 | 0=No; 1=Yes |
| q41_r | Does the public transit website explain how the public can become involved in ongoing transit issues for planning, prioritization and policy decisions? | 0,1 | 0=No; 1=Yes |

#### Table B.4 Rider Accommodations

| Variable name | Item content | Response Options | Coding |
| --- | --- | --- | --- |
| q217_r | Is information updated regularly to indicate if universal features for the BUS SYSTEM are in working order? | 0,1 | 0=No; 1=Yes |
| q227_r | Is information updated regularly to indicate if universal features for the LIGHT RAIL SYSTEM are in working order | 0,1 | 0=No; 1=Yes |
| q237_r | Is information updated regularly to indicate if universal features for the HEAVY RAIL SYSTEM are in working order? | 0,1 | 0=No; 1=Yes |
| q247_r | Is information updated regularly to indicate if universal features for the Other(1) SYSTEM are in working order? | 0,1 | 0=No; 1=Yes |
| q257_r | Is information updated regularly to indicate if universal features for the Other(2) SYSTEM are in working order? | 0,1 | 0=No; 1=Yes |
| q267_r | Is information updated regularly to indicate if universal features for the Other(3) SYSTEM are in working order? | 0,1 | 0=No; 1=Yes |
| q31_r | Does the website provide information about how to request reasonable accommodations/modifications to policies and procedures? | 0,1 | 0=No; 1=Yes |
| q32_emai | What method is used to request reasonable accommodations/modifications to policies and procedures? Check all that apply. | 0,1 | 0=No Email; 1=Email |
| q32_iper | What method is used to request reasonable accommodations/modifications to policies and procedures? Check all that apply. | 0,1 | 0=No In-Person; 1=In-Person |
| q32_mail | What method is used to request reasonable accommodations/modifications to policies and procedures? Check all that apply. | 0,1 | 0=No Mail; 1=Mail |
| q32_noinfo | What method is used to request reasonable accommodations/modifications to policies and procedures? Check all that apply. | 0,1 | 0=Information Available; 1=No Information Available |
| q32_oth | What method is used to request reasonable accommodations/modifications to policies and procedures? Check all that apply. | 0,1 | 0=No Other; 1=Other |
| q32_tele | What method is used to request reasonable accommodations/modifications to policies and procedures? Check all that apply. | 0,1 | 0=No Telephone; 1=Telephone |
| q32_webf | What method is used to request reasonable accommodations/modifications to policies and procedures? Check all that apply. | 0,1 | 0=No Web-based Form; 1=Web-based Form |
| q34_r | Telephone is used to request reasonable accommodations/modifications, is text telephone (TTY/TDD) available? | 0,1 | 0=No; 1=Yes |

#### Table B.5 Paratransit Services

| Variable name | Item content | Response Options | Coding |
| --- | --- | --- | --- |
| q271_r | Are details about PARATRANSIT (e.g. eligibility, hours of operation) available on the website? | 0,1 | 0=No; 1=Yes |
| q272_r | Is the eligibility criteria for the PARATRANSIT services clearly defined on the website? | 0,1 | 0=No; 1=Yes |
| q273_1_a | What are the hours of operation for the PARATRANSIT service? Check all that apply. - Monday | 0,1 | 0=Not Available Monday Afternoon; 1=Available Monday Afternoon |
| q273_1_e | What are the hours of operation for the PARATRANSIT service? Check all that apply. - Monday | 0,1 | 0=Not Available Monday Evening; 1=Available Monday Evening |
| q273_1_em | What are the hours of operation for the PARATRANSIT service? Check all that apply. - Monday | 0,1 | 0=Not Available Monday Early Morning; 1=Available Monday Early Morning |
| q273_1_m | What are the hours of operation for the PARATRANSIT service? Check all that apply. - Monday | 0,1 | 0=Not Available Monday Morning; 1=Available Monday Morning |
| q273_2_a | What are the hours of operation for the PARATRANSIT service? Check all that apply. - Tuesday | 0,1 | 0=Not Available Tuesday Afternoon; 1=Available Tuesday Afternoon |
| q273_2_e | What are the hours of operation for the PARATRANSIT service? Check all that apply. - Tuesday | 0,1 | 0=Not Available Tuesday Evening; 1=Available Tuesday Evening |
| q273_2_em | What are the hours of operation for the PARATRANSIT service? Check all that apply. - Tuesday | 0,1 | 0=Not Available Tuesday Early Morning; 1=Available Tuesday Early Morning |
| q273_2_m | What are the hours of operation for the PARATRANSIT service? Check all that apply. - Tuesday | 0,1 | 0=Not Available Tuesday Morning; 1=Available Tuesday Morning |
| q273_3_a | What are the hours of operation for the PARATRANSIT service? Check all that apply. - Wednesday | 0,1 | 0=Not Available Wednesday Afternoon; 1=Available Wednesday Afternoon |
| q273_3_e | What are the hours of operation for the PARATRANSIT service? Check all that apply. - Wednesday | 0,1 | 0=Not Available Wednesday Evening; 1=Available Wednesday Evening |
| q273_3_em | What are the hours of operation for the PARATRANSIT service? Check all that apply. - Wednesday | 0,1 | 0=Not Available Wednesday Early Morning; 1=Available Wednesday Early Morning |
| q273_3_m | What are the hours of operation for the PARATRANSIT service? Check all that apply. - Wednesday | 0,1 | 0=Not Available Wednesday Morning; 1=Available Wednesday Morning |
| q273_4_a | What are the hours of operation for the PARATRANSIT service? Check all that apply. - Thursday | 0,1 | 0=Not Available Thursday Afternoon; 1=Available Thursday Afternoon |
| q273_4_e | What are the hours of operation for the PARATRANSIT service? Check all that apply. - Thursday | 0,1 | 0=Not Available Thursday Evening; 1=Available Thursday Evening |
| q273_4_em | What are the hours of operation for the PARATRANSIT service? Check all that apply. - Thursday | 0,1 | 0=Not Available Thursday Early Morning; 1=Available Thursday Early Morning |
| q273_4_m | What are the hours of operation for the PARATRANSIT service? Check all that apply. - Thursday | 0,1 | 0=Not Available Thursday Morning; 1=Available Thursday Morning |
| q273_5_a | What are the hours of operation for the PARATRANSIT service? Check all that apply. - Friday | 0,1 | 0=Not Available Friday Afternoon; 1=Available Friday Afternoon |
| q273_5_e | What are the hours of operation for the PARATRANSIT service? Check all that apply. - Friday | 0,1 | 0=Not Available Friday Evening; 1=Available Friday Evening |
| q273_5_em | What are the hours of operation for the PARATRANSIT service? Check all that apply. - Friday | 0,1 | 0=Not Available Friday Early Morning; 1=Available Friday Early Morning |
| q273_5_m | What are the hours of operation for the PARATRANSIT service? Check all that apply. - Friday | 0,1 | 0=Not Available Friday Morning; 1=Available Friday Morning |
| q273_6_a | What are the hours of operation for the PARATRANSIT service? Check all that apply. - Saturday | 0,1 | 0=Not Available Saturday Afternoon; 1=Available Saturday Afternoon |
| q273_6_e | What are the hours of operation for the PARATRANSIT service? Check all that apply. - Saturday | 0,1 | 0=Not Available Saturday Evening; 1=Available Saturday Evening |
| q273_6_em | What are the hours of operation for the PARATRANSIT service? Check all that apply. - Saturday | 0,1 | 0=Not Available Saturday Early Morning; 1=Available Saturday Early Morning |
| q273_6_m | What are the hours of operation for the PARATRANSIT service? Check all that apply. - Saturday | 0,1 | 0=Not Available Saturday Morning; 1=Available Saturday Morning |
| q273_7_a | What are the hours of operation for the PARATRANSIT service? Check all that apply. - Sunday | 0,1 | 0=Not Available Sunday Afternoon; 1=Available Sunday Afternoon |
| q273_7_e | What are the hours of operation for the PARATRANSIT service? Check all that apply. - Sunday | 0,1 | 0=Not Available Sunday Evening; 1=Available Sunday Evening |
| q273_7_em | What are the hours of operation for the PARATRANSIT service? Check all that apply. - Sunday | 0,1 | 0=Not Available Sunday Early Morning; 1=Available Sunday Early Morning |
| q273_7_m | What are the hours of operation for the PARATRANSIT service? Check all that apply. - Sunday | 0,1 | 0=Not Available Sunday Morning; 1=Available Sunday Morning |
| q273_8_m | What are the hours of operation for the PARATRANSIT service? Check all that apply. - Hours not listed | 0,1 | 0=Hours of Operation Listed; 1=Hours of Operation Not Listed |
| q273_9_FRS | What are the hours of operation for the PARATRANSIT service? Check all that apply. - Same as fixed route | 0,1 | 0=Hours of Operation not the Same as Fixed Route; 1=Hours of Operation the Same as Fixed Route |
| q274_r | How far in advance do you need to make a reservation for the PARATRANSIT service? | 0,1,2 | 0=Not Listed or More than 7 Days; 1=3-7 Days; 2= < 2 Days |

#### Table B.6 Website Accessibility

| Variable name | Item content | Response Options | Coding |
| --- | --- | --- | --- |
| q14 | Please copy and paste the public transit landing page URL. | Open text | n/a |
| q35 | A web-based form is used to request reasonable accommodations/modifications, please copy and paste the webpage URL. | Open text | n/a |
| q311 | If web-based form used to submit a complaint, please copy and paste the webpage URL. | Open text | n/a |
| q51 | Please copy and paste the website URL for accessibility details. | Open text | n/a |
| q52 | Please copy and paste the website URL for maps of public transit system. If not available write "n/a" routes | Open text | n/a |
| q53 | Please copy and paste the website URL for real-time transit information for system. If not available write "n/a" | Open text | n/a |
