## Supplement C Score Generation for "TRansit ACessibility Tool (TRACT): Developing a novel scoring system for public transportation system accessibility"

### Appendix C. Data dictionary with item level recoding, dimension creation, and overall accessibility information score generation.

#### Table C.1 Facility Accessibility

| Generated Variable | Variable Content | Scoring |
| --- | --- | --- |
| fac_acpr | Facility accessible parking | (q211_acpr q221_acpr q231_acpr q241_acpr q251_acpr q261_acpr)/ # of available modes |
| fac_acrr | Facility accessible restrooms | (q211_acrr q221_acrr q231_acrr q241_acrr q251_acrr q261_acrr)/ / # of available modes |
| fac_altf | Facility offers information in alternative formats | (q211_altf q221_altf q231_altf q241_altf q251_altf q261_altf) / # of available modes |
| fac_dwar | Facilty has detectable warnings | (q211_dwar q221_dwar q231_dwar q241_dwar q251_dwar q261_dwar) / # of available modes |
| fac_elev | Facility has elevators | (q221_elev q231_elev q241_elev q251_elev q261_elev) / # of available modes |
| fac_esca | Facility has escalators | (q221_esca q231_esca q241_esca q251_esca q261_esca) / # of available modes |
| fac_fltr | Facility has front line staff training on equity and inclusion | (q211_fltr q221_fltr q231_fltr q241_fltr q251_fltr q261_fltr) / # of available modes |
| fac_gate | Facility has wider fare gates | (q211_gate q221_gate q231_gate q241_gate q251_gate q261_gate)/ # of available modes |
| fac_hast | Facility has hearing assistance system | (q211_hast q221_hast q231_hast q241_hast q251_hast q261_hast) / # of available modes |
| fac_hlop | Facility has hearing loops | (q211_hlop q221_hlop q231_hlop q241_hlop q251_hlop q261_hlop)/ # of available modes |
| fac_illu | Facility illuminates surfaces | (q211_illu q221_illu q231_illu q241_illu q251_illu q261_illu) / # of available modes |
| fac_lvbr | Facility offers level boarding | (q211_lvbr q221_lvbr q231_lvbr q241_lvbr q251_lvbr q261_lvbr) / # of available modes |
| fac_ramp | Facility has ramps available | (q211_ramp q221_ramp q231_ramp q241_ramp q251_ramp q261_ramp) / # of available modes |
| fac_slip | Facility has slip-resistant surfaces | (q211_slip q221_slip q231_slip q241_slip q251_slip q261_slip) / # of available modes |
| fac_staa | Facility has audio announcements | (q221_staa q231_staa q241_staa q251_staa q261_staa) / # of available modes |
| fac_stvi | Facility has visual information | (q221_stvi q231_stvi q241_stvi q251_stvi q261_stvi) / # of available modes |
| fac_tick | Facility sells tickets via a kiosk | (q211_tick q221_tick q231_tick q241_tick q251_tick q261_tick) / # of available modes |
| fac_tico | Facility sells tickets via online | (q211_tico q221_tico q231_tico q241_tico q251_tico q261_tico) / # of available modes |
| fac_ticp | Facility sells tickets via in-person | (q211_ticp q221_ticp q231_ticp q241_ticp q251_ticp q261_ticp) / # of available modes |
| fac_ttpg | Facility offers a travel training program | (q211_ttpg q221_ttpg q231_ttpg q241_ttpg q251_ttpg q261_ttpg) / # of available modes |
| fac_wayf | Facility has wayfinding signs | (q211_wayf q221_wayf q231_wayf q241_wayf q251_wayf q261_wayf) / # of available modes |
| fac_upd | Information about which facilities have the universal design features is listed on the website | (q213_r q223_r q233_r q243_r q253_r q263_r) / # of available modes |
| fac_total | Facility accessibility information score | fac_acpr + + + fac_acrr + fac_altf + fac_dwar + fac_elev + fac_esca + fac_fltr + fac_gate + fac_hast + fac_hlop + fac_illu + fac_lvbr + fac_ramp + fac_slip + fac_staa + fac_stvi + fac_tick + + fac_tico + fac_ticp + fac_ttpg + fac_wayf + fac_upd |

#### Table C.2 Vehicle Accessibility

| Generated Variable | Variable Content | Scoring |
| --- | --- | --- |
| veh_dwar | Vehicles have detectable warnings onboard | (q214_dwar q224_dwar q234_dwar q244_dwar q254_dwar q264_dwar)/ # of available modes |
| veh_exin | Vehicles have external information | (q214_exin q244_exin q254_exin q264_exin) / # of available modes |
| veh_gapr | Vehicles offers gap reducers to users | (q224_gapr q234_gapr q244_gapr q254_gapr q264_gapr) / # of available modes |
| veh_lben | Vehicles have a lower body entrance | (q214_lben q244_lben q254_lben q264_lben) / # of available modes |
| veh_lift | Vehicles have a lift | (q214_lift q224_lift q234_lift q244_lift q254_lift q264_lift) / # of available modes |
| veh_oban | Vehicles have on-board audio announcements | (q214_oban q224_oban q234_oban q244_oban q254_oban q264_oban) / # of available modes |
| veh_obvi | Vehicles have on-board visual information | (q214_obvi q224_obvi q234_obvi q244_obvi q254_obvi q264_obvi) / # of available modes |
| veh_ramp | Vehicles have a ramp | (q214_ramp q224_ramp q234_ramp q244_ramp q254_ramp q264_ramp) / # of available modes |
| veh_secu | Vehicles have securement devices | (q214_secu q224_secu q234_secu q244_secu q254_secu q264_secu) / # of available modes |
| veh_vsrs | Vehicles have variation in stop request signals | (q214_vsrs q224_vsrs q234_vsrs q244_vsrs q254_vsrs q264_vsrs) / # of available modes |
| veh_upd | Information about which vehicles have the universal design features is listed on the website | (q216_r q226_r q236_r q246_r q256_r q266_r) / # of available modes |
| veh_total | Vehicle accessibility information score | veh_dwar + veh_exin + veh_gapr + veh_lben + veh_lift + veh_oban + veh_obvi + veh_ramp + veh_secu + veh_vsrs + veh_upd |

#### Table C.3 Inclusive Policies

| Generated Variable | Variable Content | Scoring |
| --- | --- | --- |
| policy_total | Inclusive policies score | q36_r + q37_r + q38_emai + q38_iper + q38_mail + q38_oth + q38_tele + q38_webf + q310_r + q312_r + q41_r + q19_r |

#### Table C.4 Rider Accommodations

| Generated Variable | Variable Content | Scoring |
| --- | --- | --- |
| q217_mean | Information is updated regularly to indicate universal design features are in working order | (q217_r q227_r q237_r q247_r q257_r q267_r) / # of available modes |
| accom_total | Rider accommodations score | q31_r + q32_emai + q32_iper + q32_mail + q32_oth + q32_tele + q32_webf + q34_r + q217_mean |

#### Table C.5 Paratransit Services

| Generated Variable | Variable Content | Scoring |
| --- | --- | --- |
| para_mon | Paratransit hours of operation on Monday | (q273_1_a q273_1_e q273_1_em q273_1_m)/4 |
| para_tue | Paratransit hours of operation on Tuesday | (q273_2_a q273_2_e q273_2_em q273_2_m)/4 |
| para_wed | Paratransit hours of operation on Wednesday | (q273_3_a q273_3_e q273_3_em q273_3_m)/4 |
| para_thr | Paratransit hours of operation on Thursday | (q273_4_a q273_4_e q273_4_em q273_4_m)/4 |
| para_fri | Paratransit hours of operation on Friday | (q273_5_a q273_5_e q273_5_em q273_5_m)/4 |
| para_sat | Paratransit hours of operation on Saturday | (q273_6_a q273_6_e q273_6_em q273_6_m)/4 |
| para_sun | Paratransit hours of operation on Sunday | (q273_7_a q273_7_e q273_7_em q273_7_m)/4 |
| para_days | Paratransit hours of operation over the course of a week | para_mon + para_tue + para_wed + para_thr + para_fri + para_sat + para_sun |
| para_hrs | Summary of paratransit hours of operation accessibility information score | 0 = hours not listed or less than or equal to 5 days a week  1 = hours same as fixed route system or 5-6 days a week  2 = operates greater than 6 days a week |
| para_total | Paratransit services score | q271_r + q272_r + para_hrs + q274_r |

#### Table C.6 Website Accessibility

| Generated Variable | Variable Content | Scoring |
| --- | --- | --- |
| website_total | Website Accessibility information score | 0 = bottom tertile  1 = middle tertile  2 = top tertile |

#### Table C.7 Overall Accessibility information score

| Generated Variable | Variable Content | Scoring |
| --- | --- | --- |
| overall_total | Overall accessibility information score | fac_total + veh_total + para_total + website_total + accom_total + policy_total |
