## Supplement D Syntax for "TRansit ACessibility Tool (TRACT): Developing a novel scoring system for public transportation system accessibility"

### Appendix D. Syntax used to create the six dimensions and overall accessibility public transportation score.

#### Facility Accessibility

egen fac_acpr = rmean(q211_acpr q221_acpr q231_acpr q241_acpr q251_acpr q261_acpr)

egen fac_acrr = rmean(q211_acrr q221_acrr q231_acrr q241_acrr q251_acrr q261_acrr)

egen fac_altf = rmean(q211_altf q221_altf q231_altf q241_altf q251_altf q261_altf)

egen fac_dwar = rmean(q211_dwar q221_dwar q231_dwar q241_dwar q251_dwar q261_dwar)

egen fac_elev = rmean(q221_elev q231_elev q241_elev q251_elev q261_elev)

egen fac_esca = rmean(q221_esca q231_esca q241_esca q251_esca q261_esca)

egen fac_fltr = rmean(q211_fltr q221_fltr q231_fltr q241_fltr q251_fltr q261_fltr)

egen fac_gate = rmean(q211_gate q221_gate q231_gate q241_gate q251_gate q261_gate)

egen fac_hast = rmean(q211_hast q221_hast q231_hast q241_hast q251_hast q261_hast)

egen fac_hlop = rmean(q211_hlop q221_hlop q231_hlop q241_hlop q251_hlop q261_hlop)

egen fac_illu = rmean(q211_illu q221_illu q231_illu q241_illu q251_illu q261_illu)

egen fac_lvbr = rmean(q211_lvbr q221_lvbr q231_lvbr q241_lvbr q251_lvbr q261_lvbr)

egen fac_ramp = rmean(q211_ramp q221_ramp q231_ramp q241_ramp q251_ramp q261_ramp)

egen fac_slip = rmean(q211_slip q221_slip q231_slip q241_slip q251_slip q261_slip)

egen fac_staa = rmean(q221_staa q231_staa q241_staa q251_staa q261_staa)

egen fac_stvi = rmean(q221_stvi q231_stvi q241_stvi q251_stvi q261_stvi)

egen fac_tick = rmean(q211_tick q221_tick q231_tick q241_tick q251_tick q261_tick)

egen fac_tico = rmean(q211_tico q221_tico q231_tico q241_tico q251_tico q261_tico)

egen fac_ticp = rmean(q211_ticp q221_ticp q231_ticp q241_ticp q251_ticp q261_ticp)

egen fac_ttpg = rmean(q211_ttpg q221_ttpg q231_ttpg q241_ttpg q251_ttpg q261_ttpg)

egen fac_wayf = rmean(q211_wayf q221_wayf q231_wayf q241_wayf q251_wayf q261_wayf)

egen fac_upd = rmean(q213_r q223_r q233_r q243_r q253_r q263_r)

egen fac_total = rowtotal(fac_acpr fac_acrr fac_altf fac_dwar fac_elev fac_esca fac_fltr fac_gate fac_hast fac_hlop fac_illu fac_lvbr fac_ramp fac_slip fac_staa fac_stvi fac_tick fac_tico fac_ticp fac_ttpg fac_wayf fac_upd)

#### Vehicle Accessibility

egen veh_dwar = rmean(q214_dwar q224_dwar q234_dwar q244_dwar q254_dwar q264_dwar)

egen veh_exin = rmean(q214_exin q244_exin q254_exin q264_exin)

egen veh_gapr = rmean(q224_gapr q234_gapr q244_gapr q254_gapr q264_gapr)

egen veh_lben = rmean(q214_lben q244_lben q254_lben q264_lben)

egen veh_lift = rmean(q214_lift q224_lift q234_lift q244_lift q254_lift q264_lift)

egen veh_oban = rmean(q214_oban q224_oban q234_oban q244_oban q254_oban q264_oban)

egen veh_obvi = rmean(q214_obvi q224_obvi q234_obvi q244_obvi q254_obvi q264_obvi)

egen veh_ramp = rmean(q214_ramp q224_ramp q234_ramp q244_ramp q254_ramp q264_ramp)

egen veh_secu = rmean(q214_secu q224_secu q234_secu q244_secu q254_secu q264_secu)

egen veh_vsrs = rmean(q214_vsrs q224_vsrs q234_vsrs q244_vsrs q254_vsrs q264_vsrs)

egen veh_upd = rmean(q216_r q226_r q236_r q246_r q256_r q266_r)

egen veh_total = rowtotal(veh_dwar veh_exin veh_gapr veh_lben veh_lift veh_oban veh_obvi veh_ramp veh_secu veh_vsrs veh_upd)

#### Inclusive Policies

egen policy_total = rowtotal(q36_r q37_r q38_emai q38_iper q38_mail q38_oth q38_tele q38_webf q310_r q312_r q41_r q19_r)

#### Rider Accommodations

egen q217_mean = rmean(q217_r q227_r q237_r q247_r q257_r q267_r)

egen accom_total = rowtotal(q31_r q32_emai q32_iper q32_mail q32_oth q32_tele q32_webf q34_r q217_mean)

#### Paratransit Services

```
egen para_mon = rmean(q273_1_a q273_1_e q273_1_em q273_1_m)
egen para_tue = rmean(q273_2_a q273_2_e q273_2_em q273_2_m)
egen para_wed = rmean(q273_3_a q273_3_e q273_3_em q273_3_m)
egen para_thr = rmean(q273_4_a q273_4_e q273_4_em q273_4_m)
egen para_fri = rmean(q273_5_a q273_5_e q273_5_em q273_5_m)
egen para_sat = rmean(q273_6_a q273_6_e q273_6_em q273_6_m)
egen para_sun = rmean(q273_7_a q273_7_e q273_7_em q273_7_m)

egen para_days = rowtotal(para_mon para_tue para_wed para_thr para_fri para_sat para_sun)
gen para_hrs = .
	replace para_hrs = 0 if q273_8_m == 1 | (para_days > 0 & para_days <= 5)
	replace para_hrs = 1 if q273_9_FRS == 1 | (para_days > 5 & para_days <= 6)
	replace para_hrs = 2 if para_days > 6

egen para_total = rowtotal(q271_r q272_r para_hrs q274_r)
```

#### Website Accessibility

```
gen website_total = .
	replace website_total = 0 if webaim == "lowest tertile"
	replace website_total = 1 if webaim == "middle tertile"
	replace website_total = 2 if webaim == "highest tertile"
	label define website_total 0 "Worst website access" 1 "Middle website access" 2 "Best website access"
	label var website_total website_total
```

#### Overall Accessibility information score

egen overall_total = rowtotal(fac_total veh_total para_total website_total accom_total policy_total)
