## Supplement E Reliability for "TRansit ACessibility Tool (TRACT): Developing a novel scoring system for public transportation system accessibility"

### Appendix E. Reliability statistics for overall public transportation accessibility information score, dimensions of accessibility, and individual items.

Insert

#### Table E.1 Agreement between coders on the overall transportation accessibility information score and dimensions of accessibility

|  | ICC | 95% CI |
| --- | --- | --- |
| Overall Accessibility | 0.47 | (0.0, 0.8) |
| Facility Accessibility | 0.17 | (-0.1, 0.5) |
| Vehicle Accessibility | 0.51 | (0.0, 0.8) |
| Inclusive Policies | 0.30 | (-0.1, 0.6) |
| Rider Accommodations | 0.70 | (0.4, 0.9) |
| Paratransit Services | 0.63 | (0.3, 0.8) |

#### Table E.2 Prevalence, percent agreement, and Cohen’s kappa values for 140 TRansit ACessibility Tool (TRACT) items with prevalence greater than 0.1

| Item | N | Average Prevalence | Percent Agreement  (%) | Cohen's kappa  κ (95% CI) |
| --- | --- | --- | --- | --- |
| Facility Accessibility Items | | | | |
| q211_acpr | 23 | 0.1 | 78.3 | -0.1 (-0.4, 0.2) |
| q211_altf | 23 | 0.2 | 82.6 | 0.4 (0.1, 0.8) |
| q211_fltr | 23 | 0.3 | 73.9 | 0.4 (0.0, 0.8) |
| q211_hast | 23 | 0.1 | 91.3 | 0.5 (0.1, 0.8) |
| q211_tick | 23 | 0.7 | 56.5 | 0.1 (-0.3, 0.5) |
| q211_tico | 23 | 1.0 | 73.9 | 0.2 (-0.1, 0.4) |
| q211_ticp | 23 | 1.0 | 65.2 | 0.1 (-0.2, 0.4) |
| q211_ttpg | 23 | 0.7 | 43.5 | 0.0 (-0.3, 0.3) |
| q211_wayf | 23 | 0.2 | 78.3 | 0.2 (0.0, 0.5) |
| q213_r | 23 | 0.5 | 56.5 | 0.2 (-0.1, 0.4) |
| q221_acpr | 19 | 0.3 | 68.4 | 0.1 (-0.3, 0.5) |
| q221_altf | 19 | 0.5 | 68.4 | 0.4 (0.0, 0.8) |
| q221_dwar | 19 | 0.7 | 52.6 | 0.1 (-0.3, 0.5) |
| q221_elev | 19 | 0.5 | 84.2 | 0.7 (0.3, 1.1) |
| q221_esca | 19 | 0.3 | 84.2 | 0.6 (0.2, 1.0) |
| q221_fltr | 19 | 0.2 | 89.5 | 0.6 (0.2, 1.0) |
| q221_hast | 19 | 0.1 | 84.2 | 0.3 (-0.1, 0.8) |
| q221_lvbr | 19 | 0.7 | 68.4 | 0.4 (-0.1, 0.8) |
| q221_ramp | 19 | 0.5 | 89.5 | 0.8 (0.3, 1.2) |
| q221_staa | 19 | 0.4 | 78.9 | 0.4 (0.0, 0.8) |
| q221_stvi | 19 | 0.3 | 84.2 | 0.6 (0.2, 1.0) |
| q221_tick | 19 | 0.8 | 68.4 | 0.3 (0.0, 0.7) |
| q221_tico | 19 | 0.9 | 57.9 | 0.0 (-0.3, 0.3) |
| q221_ticp | 19 | 0.7 | 57.9 | 0.2 (-0.2, 0.6) |
| q221_ttpg | 19 | 0.7 | 47.4 | 0.2 (-0.1, 0.4) |
| q221_wayf | 19 | 0.4 | 78.9 | 0.6 (0.2, 1.0) |
| q223_r | 19 | 0.5 | 52.6 | 0.2 (-0.1, 0.5) |
| q231_acpr | 17 | 0.5 | 88.2 | 0.8 (0.3, 1.2) |
| q231_acrr | 17 | 0.1 | 82.4 | 0.3 (-0.2, 0.8) |
| q231_altf | 17 | 0.6 | 64.7 | 0.3 (-0.1, 0.8) |
| q231_dwar | 17 | 0.7 | 76.5 | 0.5 (0.0, 1.0) |
| q231_elev | 17 | 0.9 | 82.4 | 0.6 (0.1, 1.0) |
| q231_esca | 17 | 0.5 | 82.4 | 0.6 (0.2, 1.1) |
| q231_fltr | 17 | 0.3 | 88.2 | 0.7 (0.2, 1.1) |
| q231_gate | 17 | 0.2 | 94.1 | 0.0 (0.0, 0.0) |
| q231_hast | 17 | 0.3 | 64.7 | 0.0 (-0.5, 0.5) |
| q231_hlop | 17 | 0.1 | 94.1 | 0.6 (0.2, 1.1) |
| q231_lvbr | 17 | 0.3 | 76.5 | 0.4 (0.0, 0.8) |
| q231_ramp | 17 | 0.4 | 88.2 | 0.7 (0.3, 1.2) |
| q231_staa | 17 | 0.4 | 76.5 | 0.5 (0.1, 0.9) |
| q231_stvi | 17 | 0.4 | 70.6 | 0.3 (-0.2, 0.7) |
| q231_tick | 17 | 0.9 | 52.9 | -0.3 (-0.7, 0.2) |
| q231_tico | 17 | 1.0 | 52.9 | -0.3 (-0.7, 0.2) |
| q231_ticp | 17 | 0.8 | 52.9 | 0.0 (-0.4, 0.5) |
| q231_ttpg | 17 | 0.6 | 35.3 | -0.1 (-0.3, 0.2) |
| q231_wayf | 17 | 0.6 | 52.9 | 0.2 (-0.1, 0.4) |
| q233_r | 16 | 0.7 | 75.0 | 0.5 (0.1, 1.0) |
| q241_acrr | 3 | 0.2 | 66.7 | 0.0 (0.0, 0.0) |
| q241_elev | 3 | 0.2 | 66.7 | 0.0 (0.0, 0.0) |
| q241_tico | 3 | 0.7 | 33.3 | 0.0 (0.0, 0.0) |
| q241_ticp | 3 | 0.8 | 33.3 | 0.0 (0.0, 0.0) |
| q241_ttpg | 3 | 0.5 | 66.7 | 0.0 (0.0, 0.0) |
| q243_r | 2 | 0.8 | 50.0 | 0.0 (0.0, 0.0) |
| Vehicle Accessibility Items | | | | |
| q214_exin | 23 | 0.6 | 60.9 | 0.2 (-0.2, 0.6) |
| q214_lben | 23 | 0.7 | 82.6 | 0.6 (0.2, 1.0) |
| q214_lift | 23 | 0.8 | 78.3 | 0.5 (0.1, 0.9) |
| q214_oban | 23 | 0.9 | 95.7 | 0.6 (0.3, 1.0) |
| q214_obvi | 23 | 0.6 | 91.3 | 0.8 (0.4, 1.2) |
| q214_oth | 23 | 0.5 | 47.8 | 0.0 (-0.2, 0.3) |
| q214_ramp | 23 | 1.0 | 91.3 | 0.0 (-0.5, 0.4) |
| q214_secu | 23 | 0.9 | 91.3 | 0.5 (0.0, 0.9) |
| q214_vsrs | 23 | 0.5 | 82.6 | 0.7 (0.3, 1.1) |
| q216_r | 23 | 0.6 | 65.2 | 0.4 (0.1, 0.7) |
| q224_dwar | 19 | 0.2 | 78.9 | 0.3 (0.0, 0.6) |
| q224_gapr | 19 | 0.3 | 78.9 | 0.2 (-0.2, 0.6) |
| q224_lift | 19 | 0.2 | 84.2 | -0.1 (-0.5, 0.3) |
| q224_oban | 19 | 0.7 | 78.9 | 0.6 (0.2, 1.0) |
| q224_obvi | 19 | 0.5 | 84.2 | 0.7 (0.3, 1.1) |
| q224_oth | 19 | 0.7 | 63.2 | 0.3 (-0.2, 0.7) |
| q224_ramp | 19 | 0.4 | 63.2 | 0.1 (-0.3, 0.6) |
| q224_secu | 19 | 0.2 | 89.5 | 0.4 (0.0, 0.9) |
| q224_vsrs | 19 | 0.2 | 68.4 | -0.1 (-0.4, 0.2) |
| q226_r | 18 | 0.5 | 44.4 | 0.1 (-0.1, 0.4) |
| q234_dwar | 17 | 0.1 | 82.4 | 0.3 (-0.2, 0.8) |
| q234_gapr | 17 | 0.3 | 88.2 | 0.5 (0.1, 0.8) |
| q234_lift | 17 | 0.2 | 94.1 | 0.8 (0.3, 1.2) |
| q234_oban | 17 | 0.8 | 88.2 | 0.7 (0.2, 1.2) |
| q234_obvi | 17 | 0.5 | 70.6 | 0.4 (0.0, 0.9) |
| q234_oth | 17 | 0.6 | 52.9 | 0.0 (-0.4, 0.4) |
| q234_ramp | 17 | 0.1 | 94.1 | 0.6 (0.2, 1.1) |
| q234_secu | 17 | 0.1 | 94.1 | 0.6 (0.2, 1.1) |
| q234_vsrs | 17 | 0.1 | 82.4 | 0.0 (0.0, 0.0) |
| q236_r | 16 | 0.4 | 50.0 | 0.1 (-0.3, 0.5) |
| q244_oban | 3 | 0.3 | 100.0 | 1.0 (-0.1, 2.1) |
| q244_obvi | 3 | 0.3 | 100.0 | 1.0 (-0.1, 2.1) |
| q244_oth | 3 | 0.3 | 66.7 | 0.0 (0.0, 0.0) |
| q246_r | 2 | 0.4 | 100.0 | 1.0 (-0.4, 2.4) |
| Inclusive Policies Items | | | | |
| q19_r | 26 | 0.8 | 80.8 | 0.5 (0.1, 0.8) |
| q37_r | 26 | 0.3 | 53.8 | 0.0 (-0.3, 0.4) |
| q38_emai | 26 | 0.5 | 69.2 | 0.3 (-0.1, 0.7) |
| q38_iper | 26 | 0.3 | 92.3 | 0.8 (0.4, 1.2) |
| q38_mail | 26 | 0.7 | 76.9 | 0.5 (0.1, 0.9) |
| q38_oth | 26 | 0.3 | 57.7 | 0.2 (-0.1, 0.5) |
| q38_tele | 26 | 1.0 | 80.8 | 0.2 (-0.1, 0.5) |
| q38_webf | 26 | 0.8 | 65.4 | 0.1 (-0.3, 0.5) |
| q41_r | 26 | 0.8 | 65.4 | 0.2 (-0.2, 0.5) |
| Rider Accommodations Items | | | | |
| q217_r | 16 | 0.3 | 81.3 | 0.6 (0.1, 1.1) |
| q227_r | 17 | 0.7 | 58.8 | 0.2 (-0.3, 0.7) |
| q237_r | 15 | 0.6 | 73.3 | 0.5 (0.0, 1.0) |
| q31_r | 26 | 1.0 | 96.2 | 0.6 (0.3, 1.0) |
| q312_r | 26 | 0.6 | 69.2 | 0.4 (0.0, 0.8) |
| q32_emai | 26 | 0.6 | 92.3 | 0.8 (0.5, 1.2) |
| q32_iper | 26 | 0.2 | 92.3 | 0.6 (0.2, 1.0) |
| q32_mail | 26 | 0.5 | 73.1 | 0.3 (-0.1, 0.7) |
| q32_oth | 26 | 0.3 | 69.2 | 0.4 (0.0, 0.8) |
| q32_tele | 26 | 0.9 | 92.3 | 0.7 (0.3, 1.1) |
| q32_webf | 26 | 0.4 | 76.9 | 0.5 (0.1, 0.9) |
| q34_r | 17 | 0.7 | 76.5 | 0.4 (-0.1, 0.8) |
| Paratransit Services Items | | | | |
| q271_r | 23 | 1.0 | 95.7 | 0.6 (0.3, 1.0) |
| q272_r | 19 | 0.8 | 63.2 | 0.1 (-0.3, 0.4) |
| q273_1_a | 26 | 0.5 | 76.9 | 0.6 (0.2, 0.9) |
| q273_1_e | 26 | 0.5 | 76.9 | 0.6 (0.2, 0.9) |
| q273_1_em | 26 | 0.5 | 73.1 | 0.4 (0.1, 0.8) |
| q273_1_m | 26 | 0.5 | 76.9 | 0.6 (0.2, 0.9) |
| q273_2_a | 26 | 0.5 | 76.9 | 0.6 (0.2, 0.9) |
| q273_2_e | 26 | 0.5 | 76.9 | 0.6 (0.2, 0.9) |
| q273_2_em | 26 | 0.5 | 73.1 | 0.4 (0.1, 0.8) |
| q273_2_m | 26 | 0.5 | 76.9 | 0.6 (0.2, 0.9) |
| q273_3_a | 26 | 0.5 | 76.9 | 0.6 (0.2, 0.9) |
| q273_3_e | 26 | 0.5 | 76.9 | 0.6 (0.2, 0.9) |
| q273_3_em | 26 | 0.5 | 73.1 | 0.4 (0.1, 0.8) |
| q273_3_m | 26 | 0.5 | 76.9 | 0.6 (0.2, 0.9) |
| q273_4_a | 26 | 0.5 | 76.9 | 0.6 (0.2, 0.9) |
| q273_4_e | 26 | 0.5 | 76.9 | 0.6 (0.2, 0.9) |
| q273_4_em | 26 | 0.5 | 73.1 | 0.4 (0.1, 0.8) |
| q273_4_m | 26 | 0.5 | 76.9 | 0.6 (0.2, 0.9) |
| q273_5_a | 26 | 0.5 | 76.9 | 0.6 (0.2, 0.9) |
| q273_5_e | 26 | 0.5 | 76.9 | 0.6 (0.2, 0.9) |
| q273_5_em | 26 | 0.5 | 73.1 | 0.4 (0.1, 0.8) |
| q273_5_m | 26 | 0.5 | 76.9 | 0.6 (0.2, 0.9) |
| q273_6_a | 26 | 0.5 | 76.9 | 0.6 (0.2, 0.9) |
| q273_6_e | 26 | 0.5 | 76.9 | 0.6 (0.2, 0.9) |
| q273_6_em | 26 | 0.4 | 73.1 | 0.4 (0.1, 0.8) |
| q273_6_m | 26 | 0.5 | 76.9 | 0.6 (0.2, 0.9) |
| q273_7_a | 26 | 0.5 | 76.9 | 0.6 (0.2, 0.9) |
| q273_7_e | 26 | 0.5 | 73.1 | 0.5 (0.1, 0.8) |
| q273_7_em | 26 | 0.4 | 73.1 | 0.4 (0.1, 0.8) |
| q273_7_m | 26 | 0.5 | 76.9 | 0.6 (0.2, 0.9) |
| q273_9_FRS | 26 | 0.4 | 84.6 | 0.5 (0.1, 0.9) |
| q274_r | 21 | 0.9 | 90.5 | 0.7 (0.4, 1.1) |

*Note:* κ Cohen’s kappa statistic, CI confidence interval.
